# Fusion of Electronic Health Records and Radiographic Images for a Multimodal Deep Learning Prediction Model of Atypical Femur Fractures

**DOI:** 10.1101/2023.07.02.23292125

**Authors:** Jörg Schilcher, Alva Nilsson, Oliver Andlid, Anders Eklund

**Affiliations:** Department of Orthopedics and Experimental and Clinical Medicine, Faculty of Health Science, Linköping University, Linköping, Sweden; Wallenberg Centre for Molecular Medicine, Linköping University, Linköping, Sweden; Department of Biomedical Engineering, Linköping University, Linköping, Sweden; Division of Statistics and Machine Learning, Department of Computer and Information Science, Linköping University, Linköping, Sweden; Center for Medical Image Science and Visualization (CMIV), Linköping University, Linköping, Sweden

**Author notes:** Corresponding author Anders Eklund.

**Keywords:** Atypical femoral fractures, multimodal, fusion, deep learning

## Abstract

Atypical femur fractures (AFF) represent a very rare type of fracture that can be difficult to discriminate radiologically from normal femur fractures (NFF). AFFs are associated with drugs that are administered to prevent osteoporosis-related fragility fractures, which are highly prevalent in the elderly population. Given that these fractures are rare and the radiologic changes are subtle currently only 7% of AFFs are correctly identified, which hinders adequate treatment for most patients with AFF. Deep learning models could be trained to classify automatically a fracture as AFF or NFF, thereby assisting radiologists in detecting these rare fractures. Historically, for this classification task, only imaging data have been used, using convolutional neural networks (CNN) or vision transformers applied to radiographs. However, to mimic situations in which all available data are used to arrive at a diagnosis, we adopted an approach of deep learning that is based on the integration of image data and tabular data (from electronic health records) for 159 patients with AFF and 914 patients with NFF. We hypothesized that the combinatorial data, compiled from all the radiology departments of 72 hospitals in Sweden and the Swedish National Patient Register, would improve classification accuracy, as compared to using only one modality. At the patient level, the area under the ROC curve (AUC) increased from 0.966 to 0.987 when using the integrated set of imaging data and seven pre-selected variables, as compared to only using imaging data. More importantly, the sensitivity increased from 0.796 to 0.903. We found a greater impact of data fusion when only a randomly selected subset of available images was used to make the image and tabular data more balanced for each patient. The AUC then increased from 0.949 to 0.984, and the sensitivity increased from 0.727 to 0.849. These AUC improvements are not large, mainly because of the already excellent performance of the CNN (AUC of 0.966) when only images are used. However, the improvement is clinically highly relevant considering the importance of accuracy in medical diagnostics. We expect an even greater effect when imaging data from a clinical workflow, comprising a more diverse set of diagnostic images, are used.

## 1 Introduction

Orthopedic trauma to the extremities is the most-common reason for visits to emergency departments worldwide (Welfare, 2020) (Kraaijvanger et al., 2015), and together with other musculoskeletal conditions such as back pain and osteoarthritis, fractures are the major cause for years lived with disability globally (Collaborators, 2021). While most musculoskeletal conditions can be treated as non-urgent, acute fractures require urgent attention to be diagnosed correctly, followed by acute treatment to decrease the risks for short- and long-term morbidity and mortality. According to the Global Burden of Diseases, Injuries, and Risk Factors Study 2019, there were 178 million new fractures worldwide during 2019, which was an increase of 33% compared to 1990 (Collaborators, 2021). This increase in fractures likely corresponds to a similar increase in the number of radiologic examinations performed, which is the main modality used in fracture diagnostics. Only approximately 10% of radiographs (Gleadhill et al., 1987) permit the detection of a fracture, which means that in order to detect the 178 million fractures, theoretically, radiographs from 1.78 billion examinations would need to be reviewed annually (Liu et al., 2022). Reviewing and reporting this high volume of diagnostic images around the clock would create a situation with a high risk for diagnostic error (Pinto et al., 2016), which ranges from 4% to around 30% depending on the study sample (Bruno et al., 2015). These errors are the sixth most-common reason for malpractice claims in the US, even though radiologists make up less than 5% of physicians in the US (Whang et al., 2013). To detect rare disease patterns, such as the Atypical Femur Fractures (AFF) described here, through analyzing this bulk information is challenging, as disease prevalence has a profound impact on diagnostic accuracy, especially with respect to sensitivity (Willis, 2012).

AFF represent a very rare disease pattern among the already rare stress fractures of the femur. Much attention has been focused on these fractures because of the paradoxical causal relationship between the most commonly used drugs to treat osteoporosis (bisphosphonates) and a dramatically increased risk for AFF (Schilcher et al., 2011) (Black et al., 2020; Dell et al., 2012; Meier et al., 2012; Schilcher et al., 2015). With a yearly incidence of 1.1–2.2 AFF per 100,000 inhabitants in Sweden (Bogl et al., 2022), AFF comprise only about 4 % of all fractures of the femoral shaft and less than 0.25% of all fractures of the femur. Since AFF are so rare, clinicians might overlook this fracture pattern among the abundance of other fractures (Harborne et al., 2016; Zdolsek et al., 2021). From the radiologic perspective, this implies that every musculoskeletal radiologist might see <1 AFF case every year. Even if the radiologic stress fracture pattern of AFF is well-defined by the American Society for Bone and Mineral Research (Shane et al., 2014; Shane et al., 2010) (Figure 1), it seems likely that the yearly incidence is too low to maintain alertness to this condition in the clinical working situation, motivating the use of automated alerting systems (Zdolsek et al., 2021).

**Figure 1.**
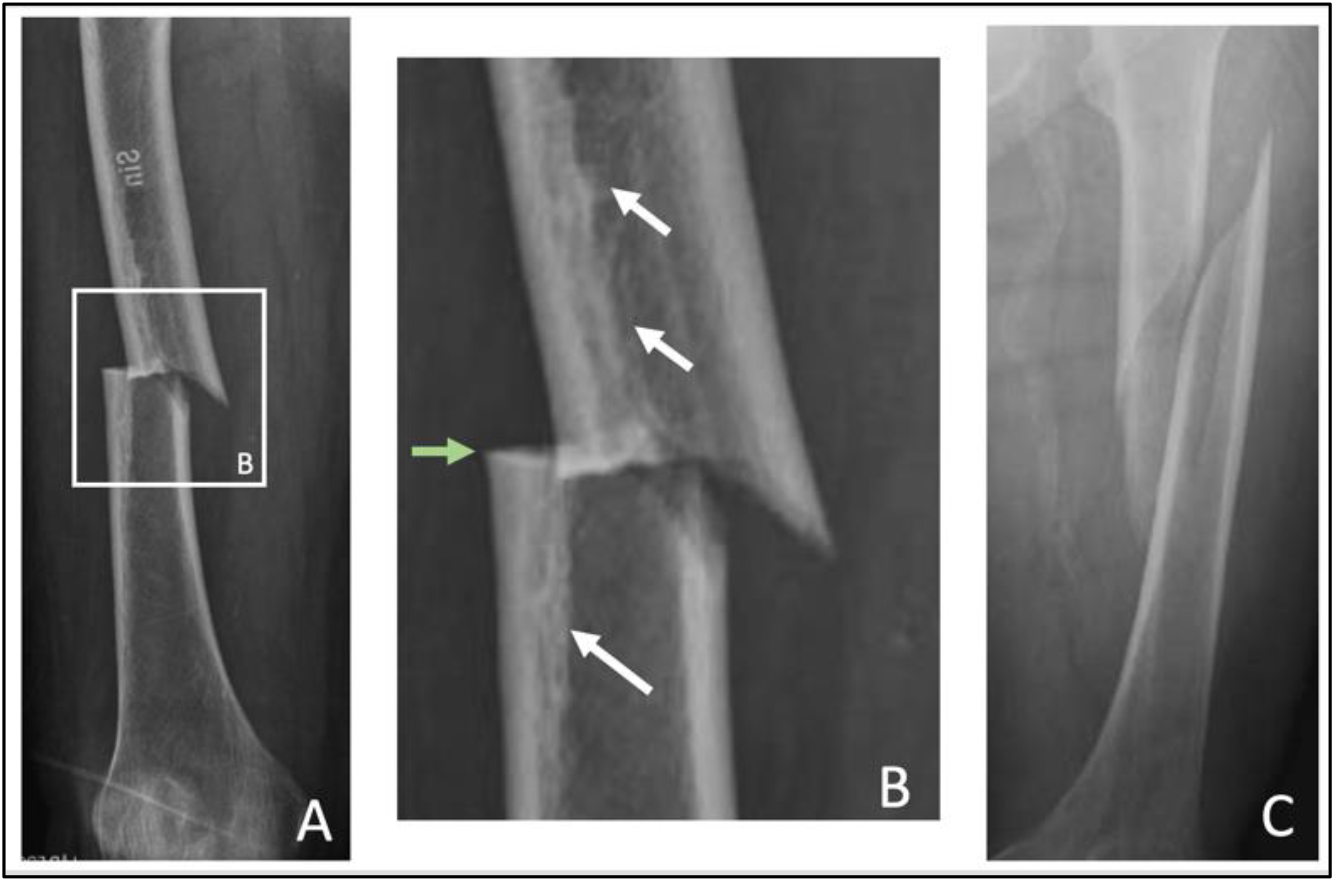
Atypical femur fracture in the diaphysis of a right femur (A). At higher magnification (B), the transverse fracture line (green arrow) and focal periosteal thickening defining the stress fracture pattern can be clearly discriminated from a normal spiral fracture (C).

A correct diagnosis is a prerequisite for successful treatment of any disease. In patients with AFF who need acute surgical treatment of their fracture, a correct diagnosis based on the first diagnostic radiographs is important because the decision regarding how to treat the fracture will be made within hours of the radiographs being acquired. A correct choice of implant for fracture fixation in AFF can minimize surgical complications (Bogl et al., 2020), and screening of the contralateral side and the timely cessation of bisphosphonate treatment can prevent the progression or development of an incomplete AFF on the contralateral side (Shane et al., 2014; Starr et al., 2018).

### 1.1 Related work on data fusion

Physicians use several sources of information, such as medical images and healthcare records, to make a diagnosis (Huang et al., 2020) because combining different types of data can increase diagnostic accuracy. In machine learning, the fusion of medical images and tabular data aims to replicate medical differential diagnostics using different sources of information. The idea of multi-modality fusion is that the different data types contribute with different independent strands of information to the AI model (Huang et al., 2020) (Acosta et al., 2022; Kline et al., 2022). Somewhat surprisingly, deep learning research on medical imaging has so far focused on imaging data, and largely ignored the wealth of clinical information available in electronic healthcare records. Currently, three different fusion techniques are used: (1) late fusion (e.g., probability fusion; PF); (2) joint fusion (e.g., feature fusion; FF); and (3) early fusion (e.g., learned feature fusion; LFF) (Huang et al., 2020). Late fusion (decision-level fusion) refers to fusing the predictions from multiple models, typically accomplished by averaging or majority voting. Probability fusion combines the predictions from an image-only network and a tabular data-only network. Joint fusion (intermediate fusion) also combines the learned features from different modalities into a single model, although the networks generating the features are not updated during the fusion training. Early fusion (feature-level fusion) combines feature vectors from imaging and tabular data into a single feature vector prior to migrating it to a dense network; the networks generating the two feature vectors are also trained during the fusion training. According to the literature, multimodal models generally outperform the corresponding single-modality models for the same tasks, although the optimal fusion technique depends on the specific application (Huang et al., 2020).

Previous work on multimodal machine learning for fractures is very limited. Yenidogan et al. have used a multimodal model to improve mortality predictions for patients with hip fractures (Yenidogan et al., 2021). Soenksen et al. (Soenksen et al., 2022) have proposed a general framework for multimodal AI for healthcare, and applied it to several different applications, such as thorax-level fractures. Recently, we have shown that a convolutional neural network (CNN) could classify AFF and normal femoral shaft fractures (NFF) with an AUC of 0.94 (Zdolsek et al., 2021). Here, we extend this work in two ways. First, the dataset is increased to cover 1,073 patients (from 373 previously). Second, we combine imaging and tabular data through a data fusion process to improve further the prediction accuracy.

## 2 Data

This work was performed using data collected from 1,124 patients in Sweden who suffered a femur fracture. The dataset was compiled from all the radiology departments of 72 hospitals in Sweden during 2011, as part of an epidemiologic research project (Schilcher et al., 2014, 2015). Images were either transferred on compact discs or transmitted electronically through an existing data transfer infrastructure provided by a picture archiving and communications systems company (Sectra AB, Sweden). After transfer, all the images were manually re-reviewed (JS) and classified as AFF (N= 172) or other fracture types (Figure 2). Among all the other fractures, NFF were identified. The diagnostic criteria for AFF were based on the ASBMR radiologic criteria (Shane et al., 2014), as characterized by a fracture that was: (1) located in the femoral shaft; (2) originated in the lateral cortex and ran perpendicular to the longitudinal axis of the femur; (3) not at all or only minimally comminuted; (4) showed a medial spike; and (5) exhibited endosteal/periosteal thickening at the fracture site. In our definition, the transverse fracture line and endosteal/periosteal thickening, both of which are pathognomonic signs of stress fractures, were compulsory features of AFF (Schilcher et al., 2013). NFF were not transverse on the lateral side. Both fracture types lacked any signs of previous surgery or malignancy. For 1,073 of the 1,124 patients, both radiographs and tabular data were available. The manual sorting process included the elimination of those images in which no fracture was present, as well as the removal of images obtained after surgery (e.g., showing an implant). Images that showed a femur fracture and no implant on one side but with an implant in the contralateral femur were retained in the dataset. A script was then used to remove exact duplicates of images. The final dataset contained a total of 4,014 radiographs (548 AFF, 3,466 NFF), with a median of three images per patient (range, 1–9) from 159 patients with AFF and 914 patients with NFF. The median image size was 2125 × 2761 pixels (range, 1,024 × 1,024 to 3,520 × 4,280). The 1,073 patients had a median age of 83 years (range, 56–102 years), and 893 were female.

**Figure 2.**
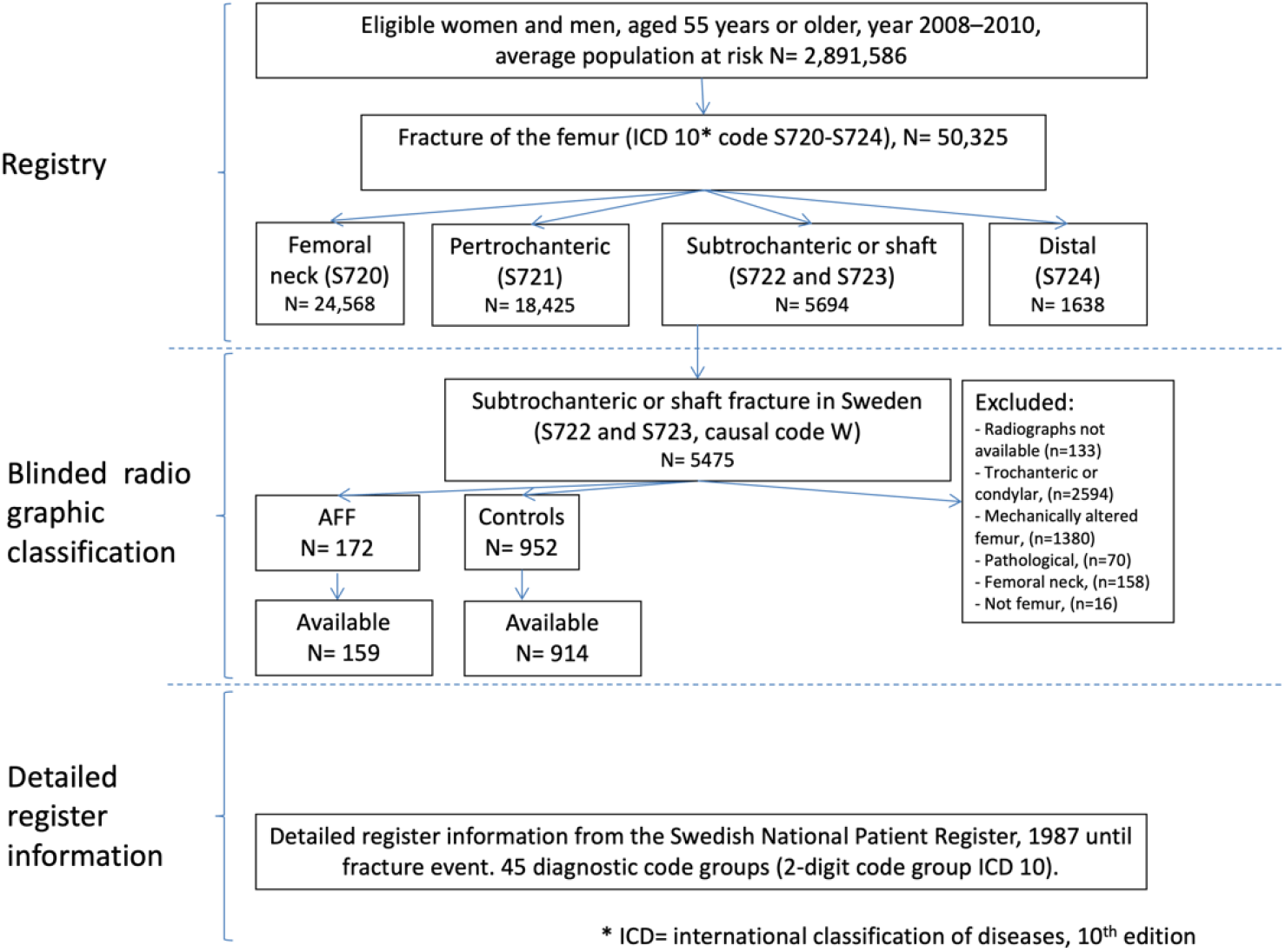
Flowchart showing the creation of the multimodal dataset using the Swedish National Patient Register. Personal numbers and tabular data were obtained from the patient register, and the images were obtained from 72 radiology departments in Sweden.

The tabular data originated from the Swedish National Patient Register^1^, which is a high-quality register of inpatient diagnoses (Ludvigsson et al., 2011). This dataset contains 45 variables for each patient (see Appendix A for a complete list of the variables) and the variables are available as binary (disease present: yes/no) or as non-binary data. In the non-binary data, the number indicates the number of days with a certain diagnosis or treatment. Seven variables are of particular interest for the present work: age, sex, osteoporosis diagnosis, rheumatoid disease, cortisone treatment, treatment with proton pump inhibitors, and bisphosphonate treatment. These variables have previously been identified as risk modifiers for AFF in regression models (Black et al., 2020; Schilcher et al., 2015). We focused on these seven variables to mimic the clinical situation in a meaningful way, deeming it unfeasible for a radiologist to keep track of 45 variables for every patient. While automatically combining radiology information with the patients’ medical charts in a clinical workflow would be challenging under Swedish law due to privacy restrictions, it might be possible in an integrated healthcare system. The dataset used in this work is unique and comprises high-quality register data and high-quality digital radiographic data for one of the largest AFF cohorts in the world.

## 3 Methods

The following sections describe the preprocessing of the data and the different prediction models. All the models were implemented in Keras 2.6 and trained on a computer with a 10-core CPU, 128 GB RAM and an Nvidia RTX 3090 graphics card with 24 GB of memory. The developed code is shared on Github at https://github.com/wanderine/AFF_fusion.

### 3.1 Preprocessing

The DICOM images (from modality computed radiography (CR) and digital x-ray imaging (DX)) were first converted to 16-bit PNG for anonymization purposes and to fit into the deep learning frameworks (not supporting the DICOM format). A large portion of the DICOM images were stored with inverted intensity, indicated in the DICOM tag “PhotometricInterpretation”. To create images with uniform intensity, all the images were converted to an intensity in the range of 0–50,000. All the images were pseudonymized using ID numbers from the tabular data. All of the images were made square using zero padding, and then down-sampled to 224 × 224 pixels, so as to fit CNN that were pre-trained on ImageNet. Down-sampling was performed using the OpenCV function cv2.resize (Bradski, 2000) with the “INTER_AREA” option.

### 3.2 Deep learning

Several different classification networks were implemented to compare the performances of different input data and different fusion approaches. The baseline networks used only imaging data or tabular data as the input. The three fusion networks (probability fusion, feature fusion and learned feature fusion) used imaging data and tabular data simultaneously. The multimodal dataset was randomly split into six equal parts, one for testing and the remaining five for cross-validation. During the training process, four parts were used for training and the fifth part was used for validation (early stopping). To avoid bias, all the radiographs from one patient were aggregated and analyzed together (Tampu et al., 2022). To ensure this patient-specific analysis of radiographs, a dictionary-object that contained several dictionaries was used, to divide randomly the patients (and all their images) into training, validation, and test groups. However, cross-validation is sensitive to the test set used. To assess more accurately the performance of each network, the process of randomization was repeated five times (each comprising 5-fold cross-validation) (Shi et al., 2010). For all the networks, binary cross-entropy was used as the loss function and Adam was used as the optimizer. Early stopping was used based on the validation loss, to prevent overfitting to the training data and restoring the best weights. Patience was set to 100 epochs. The dataset contains an unequal number of radiographs for each type of fracture (14.8% AFF). To compensate for this imbalance, proportional class weights were used during the training.

### 3.3 Classification using images only

CNN are the standard choice for image classification tasks. Inspired by the previous work on a similar but much smaller dataset (Zdolsek et al., 2021), a pre-trained ResNet50 (He et al., 2016) was selected as the image classifier for this study. As our classification task only has two classes (AFF and NFF), compared to the 1,000 in ImageNet, the last, fully connected layers were modified with inspiration from a previous study (Holste et al., 2021), and the used CNN is shown in Figure 3. Since radiographs differ substantially from the images in the ImageNet dataset, it is not sufficient to train just the last dense part of the network. Therefore, re-training was carried out from layer 39 (out of 178 layers) in the ResNet50 model.

**Figure 3.**
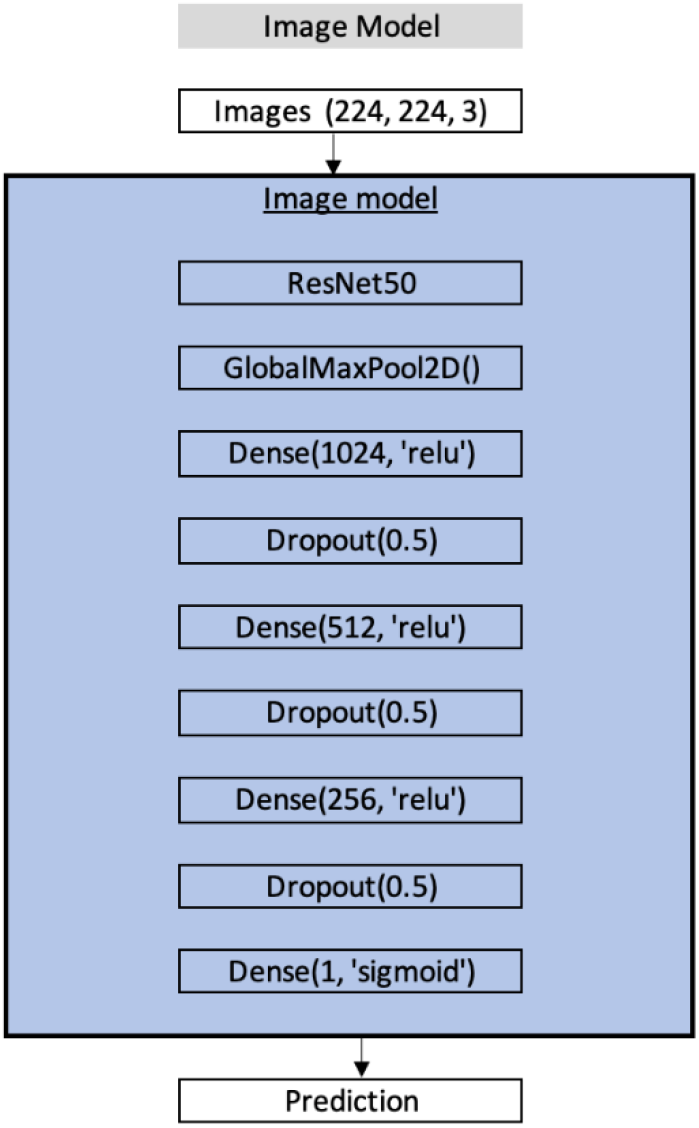
The CNN used for classification of AFF and NFF using only imaging data. The CNN is based on a pre-trained ResNet50.

On-the-fly data augmentation was used to limit overfitting, and this included random flipping (horizontal and vertical), random rotations (-180° to 180°), random translations (±10% in each direction), random zooming (±10%), and random contrast adjustment (factor of 0.3). A learning rate scheduler was used, where the learning rate started at 10^-5^ and was constant throughout the first 50 epochs. The learning rate then decreased exponentially with a factor of 10^-0.05^ per epoch, until it reached a value of 10^-6^. Thereafter, the learning rate remained constant. The learning rate was lower compared to that when training a CNN from random weights, so as not to update the pre-trained weights too quickly. Training the CNN for one fold took about 22 minutes.

### 3.4 Classification using tabular data only

The baseline model using only tabular data was based on previous research on medical conditions, such as prediction of decompensation in heart failure (Guo et al., 2020) and breast cancer classification using magnetic resonance (MR) imaging (Holste et al., 2021). Due to the low number of variables in the tabular data, this network has a low number of trainable parameters (Figure 4).

### 3.5 Classification using data fusion

The two single-modality networks were used as baseline methods and were compared with the three fusion networks. In a fusion model, it is necessary to ensure that the imaging and tabular data originate from the same patient. Therefore, the tabular data were duplicated for each available radiograph, yielding a multimodal dataset that consisted of 4,014 pairs of images and sets of tabular data variables. All the fusion networks used pre-trained baseline models for the imaging data, and to avoid bias, the same patients were used in each corresponding fold during the training of the fusion models. To ensure coherent use of patient data between the baseline and fusion forms, the weights of both baseline models (imaging and tabular data) were saved for each cross-validation fold. When training the fusion models, each cross-validation fold used the saved imaging and tabular data models corresponding to the same fold.

#### 3.5.1 Probability fusion

The architecture of the probability fusion network (which is an example of late fusion) is heavily influenced by a previous work (Holste et al., 2021) (Figure 4). In Figure 4, the blue region, illustrating the CNN, shows the baseline model using imaging data as the input. The green region, illustrating the tabular data model, shows the baseline model using tabular data as the input. As mentioned above, the weights from the corresponding folds of the pre-trained baseline models were used when a certain fold of the fusion model was trained. In this fusion method, both baseline models are running in inference mode, producing one prediction each. These two separate predictions are then concatenated and fed into a shallow neural network, to yield a final prediction. The layers shaded in yellow are trainable. A scheduler was used to control the learning rate. It started at 10^-4^ and then decreased by a factor of 10^-0.01^ per epoch, until it reached the value of 10^-5^. Training the network for one fold took about 7 minutes.

**Figure 4.**
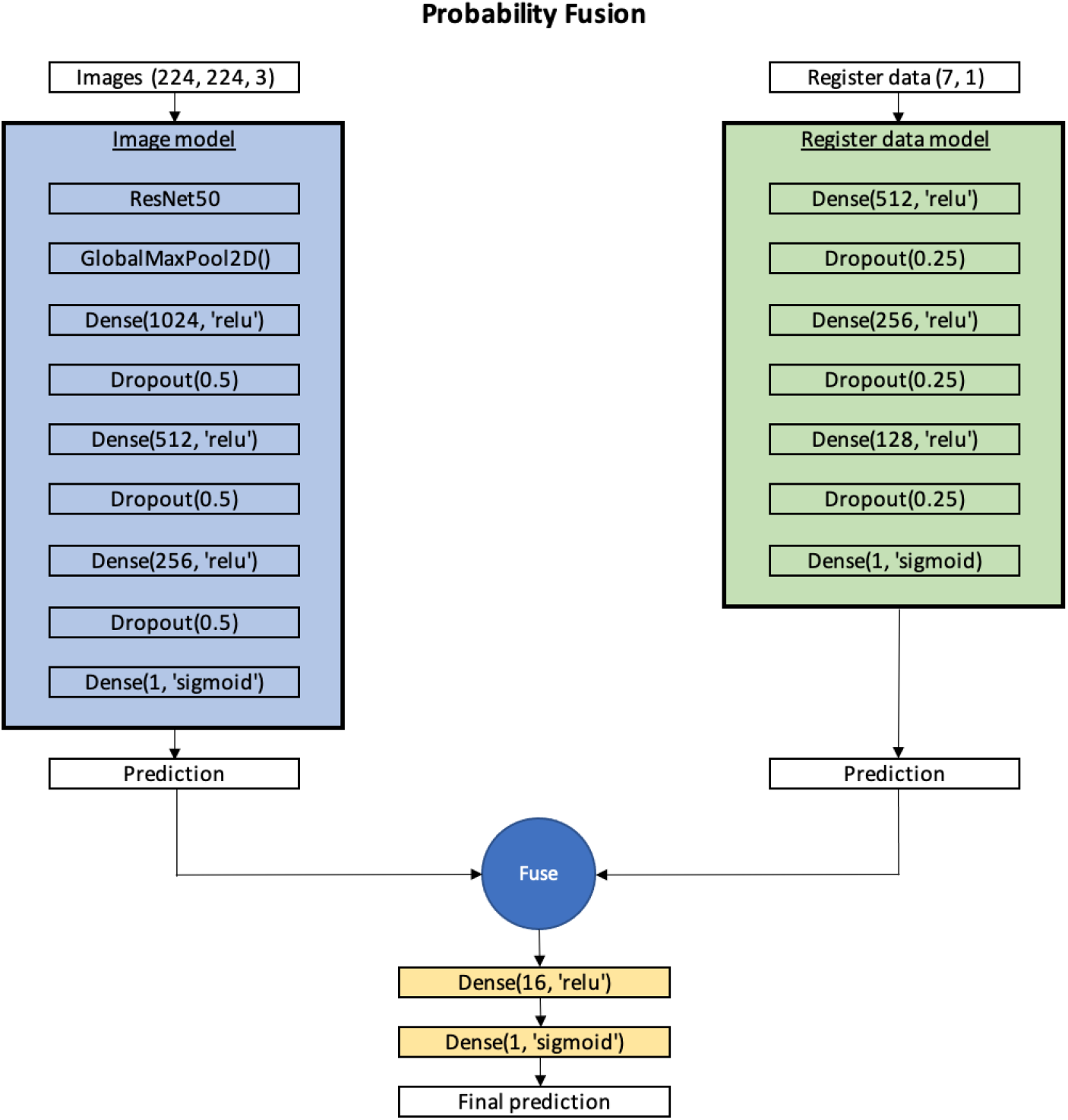
Diagram of the probability fusion architecture. The layers shaded in yellow are updated during the training. The trained baseline models are running in inference mode, and each model returns a probability for AFF. On the left is the image model, and on the right is the tabular data model. The two probabilities are then combined and fed into a trainable, small, dense network.

#### 3.5.2 Feature fusion

The architecture of the feature fusion network (which is an example of early fusion) concatenates a 2,048-sized feature vector, obtained from a radiograph, with a vector that comprises the 2–45 variables from the tabular data for the same patient (Holste et al., 2021). Prior to concatenation, both feature vectors were normalized with respect to the mean and standard deviation for each of the feature vectors. The pre-trained ResNet50 CNN was the same as that used in the probability fusion method. However, in this fusion method, a feature vector of size 2,048 is extracted from the model, instead of a scalar prediction. As in the case of the probability fusion, both baseline models were running in inference mode (no weights were updated). The combined vector was then fed into a shallow neural network, to generate a final prediction. The structure of the network is shown in Figure 5, where the layers shaded in yellow are trainable. The learning rate had a constant value of 10^-4^. Training the network for one fold took about 9 minutes.

**Figure 5.**
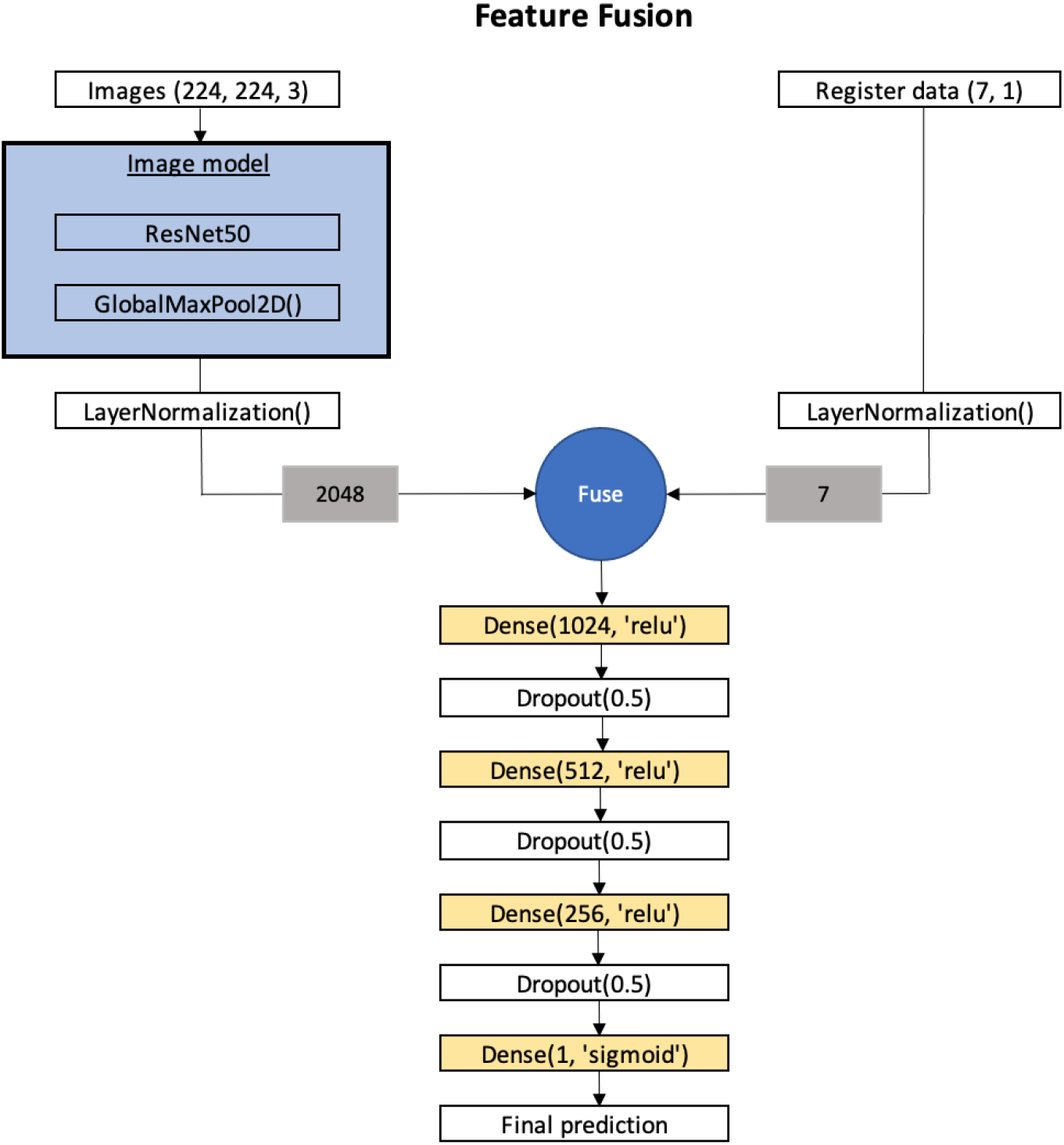
Diagram of the feature fusion architecture. The layers shaded in yellow are trainable and the baseline models are running in inference mode. The feature vectors from the baseline models are concatenated into a single-feature vector, which is then fed into a trainable dense network.

#### 3.5.3 Learned feature fusion

The learned feature fusion architecture is an example of joint fusion (Holste et al., 2021). This model also used the ResNet50 as the image model. In addition, three trainable, fully connected layers were added to derive a feature vector with size of 128. This feature vector was concatenated with another feature vector, also of size 128, which was derived by the tabular data model. This model was based on the same model as that used for the probability fusion, but instead of producing a scalar prediction it learned a feature vector representation. Unlike the other two fusion approaches, this approach combines features that were learned and updated during the training. As in the feature fusion model, the extracted features from an image and the tabular data variables were normalized. The concatenated feature vector was fed into a shallow neural network to produce a final prediction (Figure 6). A constant learning rate of 10^-4^ was used during the training of this model. Training the network for one fold took about 11 minutes.

**Figure 6.**
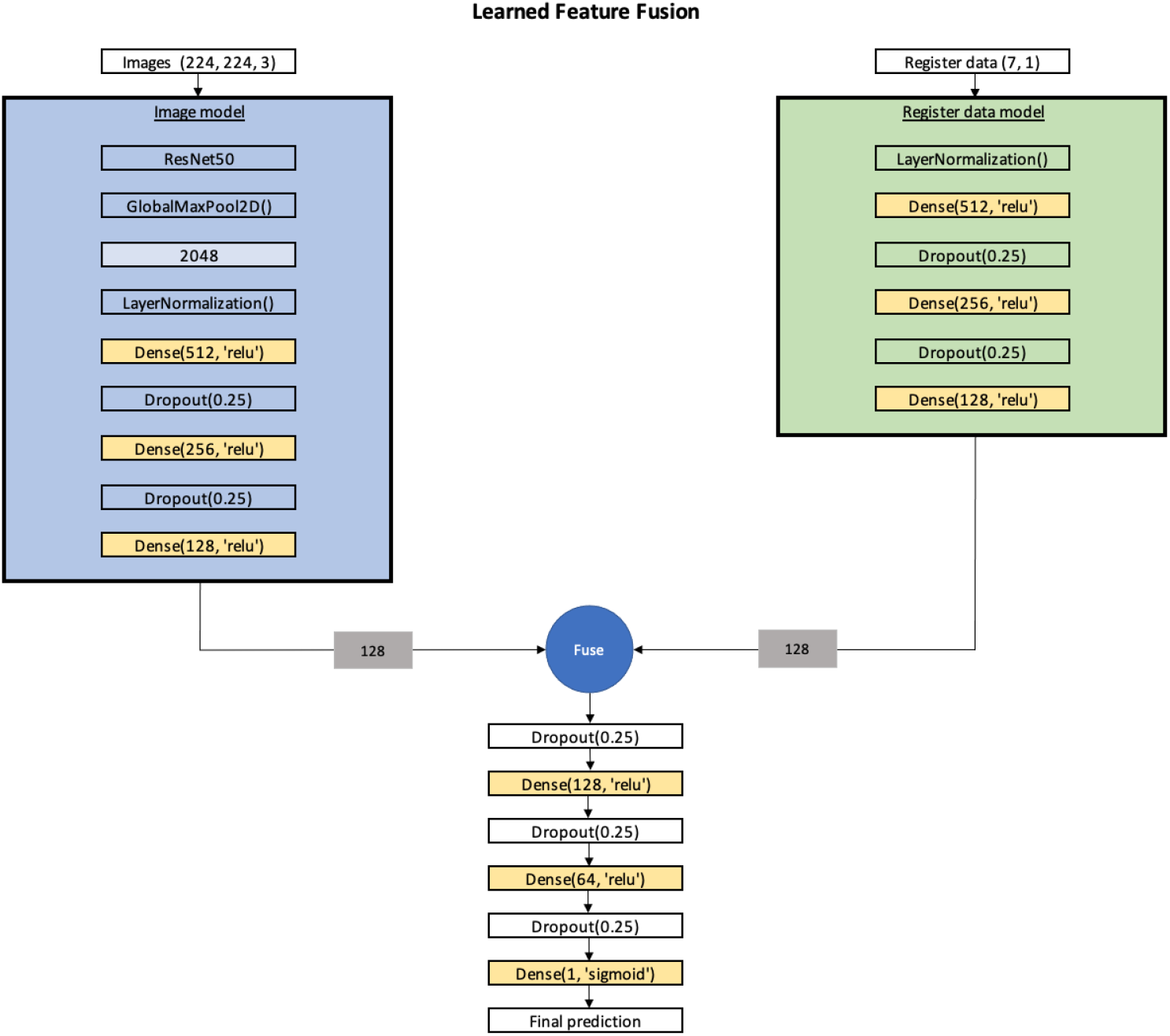
Diagram of the learned feature fusion architecture. The layers shaded in yellow are trainable. The learned feature vectors from the image network and the tabular data network are combined into a single vector and fed into a dense network. The baseline models start from the pre-trained weights and are further updated during the joint training.

### 3.6 Post-processing

The CNN output was one prediction per image. Most of the patients had more than one x-ray image available (range, 1–9), which led to multiple output predictions. However, the desired output was a single prediction per patient, i.e., the probability of a patient being diagnosed as AFF or NFF. Therefore, the post-processing of all the models with images as inputs needed to consider all the predictions for a certain patient, aggregated into a single prediction. Our approach was to calculate the mean of all the predictions (float values between 0 and 1), and to convert this mean into a binary classification.

### 3.7 Evaluation

The different networks were compared using the area under the ROC curve (AUC), sensitivity, specificity, and Matthew’s correlation coefficient (MCC) (Matthews, 1975). MCC has been shown to be superior for unbalanced cases such as ours (Chicco and Jurman, 2020), as it considers true positives, true negatives, false positives, and false negatives together to calculate a value between –1 and 1. All the metrics were calculated using functions in scikit-learn (Pedregosa et al., 2011). Each fusion training was performed in three different ways: (1) with only two tabular variables (age, sex); (2) with the seven pre-chosen variables; (3) and with all 45 variables. Each training with seven or all variables was performed with the binary as well as the non-binary versions of the variables.

As each patient had 1–9 images with more than 4 million pixels on average (about 50,000 when down-sampled to 224 × 224 pixels), there was a significant imbalance between the imaging data (4–36 million pixels, or 50,000–450,000 when down-sampled) and the tabular data (2–45 variables) for the average patient. Therefore, we repeated the training of the different models with a maximum of one and two randomly selected images from each patient, to render the two modalities more balanced.

## 4 Results

### 4.1 Fusion using all available images

Figures 7–9 show the comparisons of the different networks reported on the patient level (not the image level). The figures show the mean values and standard deviations for all the metrics over the 25 training sessions (five times repeated 5-fold CV). In the text below, we refer to the mean metrics calculated over the 25 trainings. Table 1 summarizes the most important findings when using the seven pre-selected tabular variables.

**Table 1.**
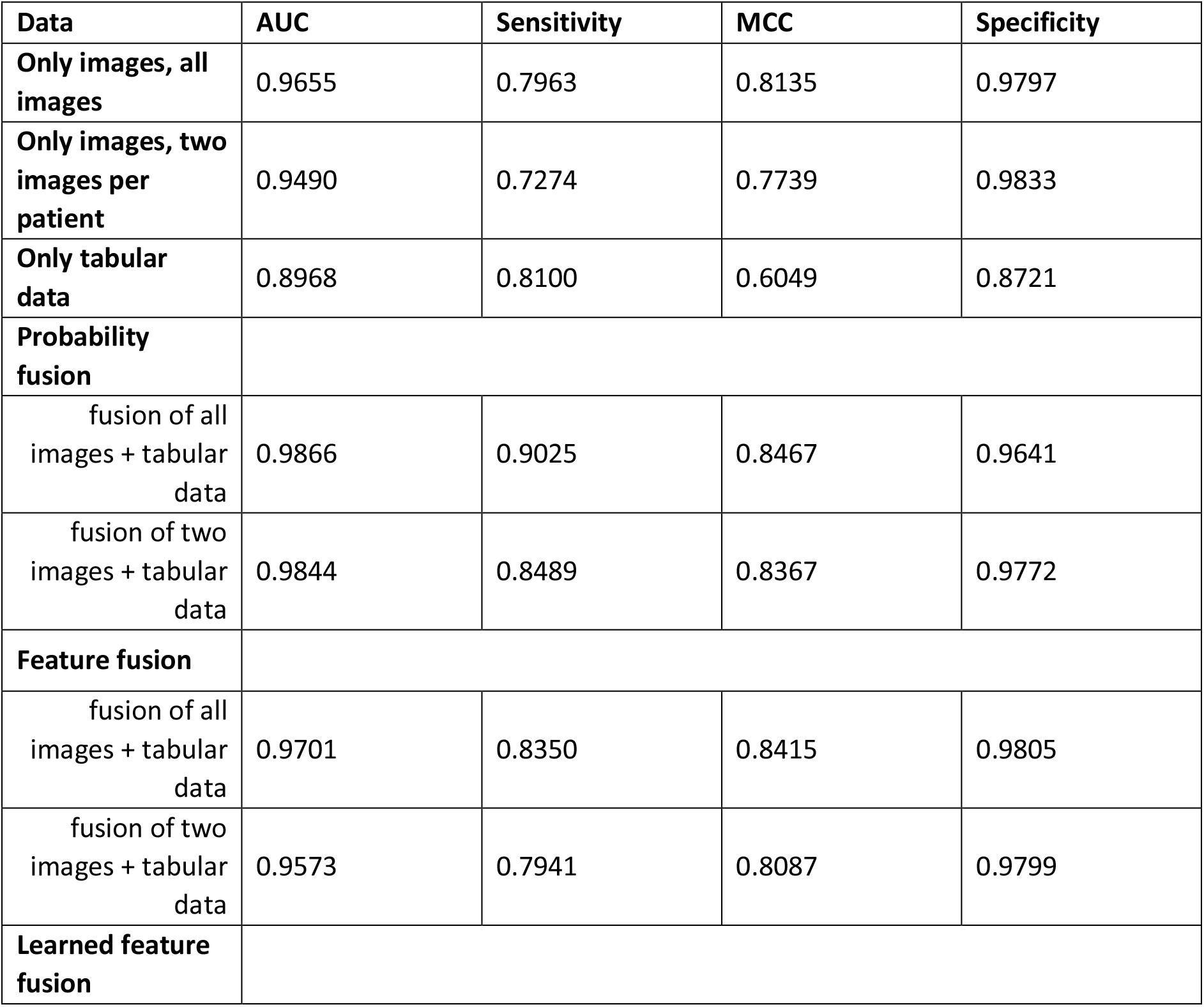
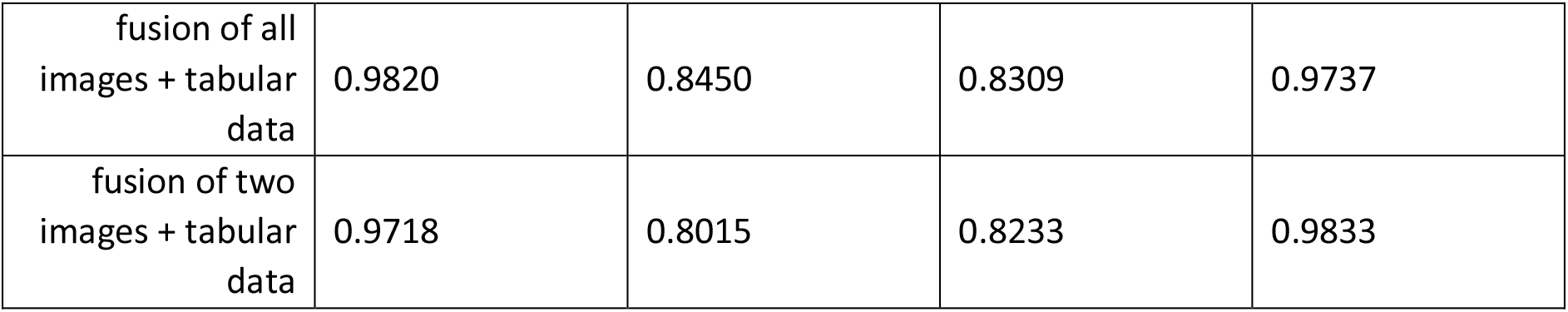
Summary of the most important results, using seven non-binary tabular variables. Each value represents the mean of the 25 trainings. Data fusion improves the predictions, especially when the two modalities are more balanced (fewer images per patient).

**Figure 7.**
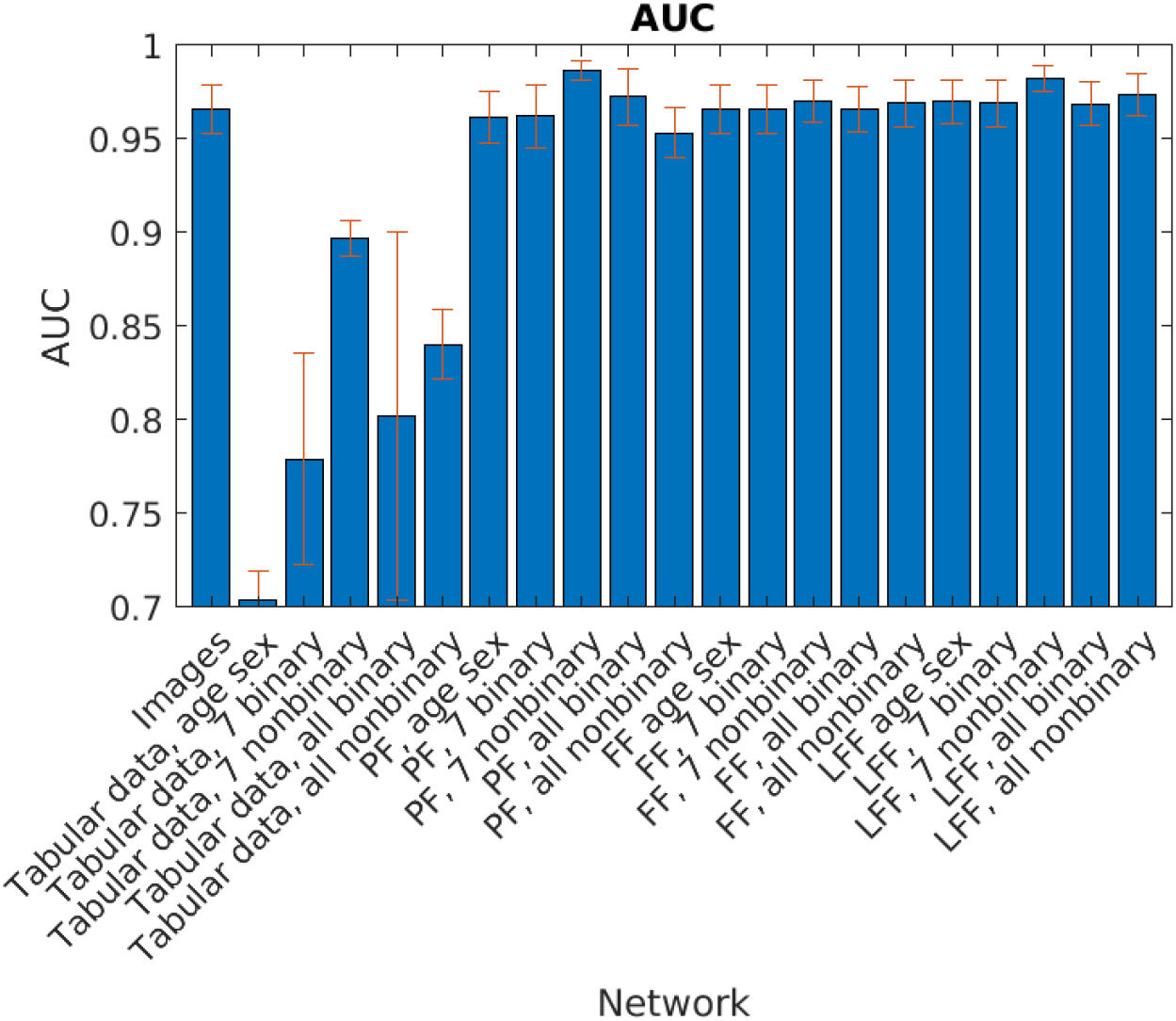
AUC values for the different networks when using all the available radiographic images per patient. PF, Probability fusion; FF, feature fusion; LFF, learned feature fusion. The error bars represent standard deviations.

**Figure 8.**
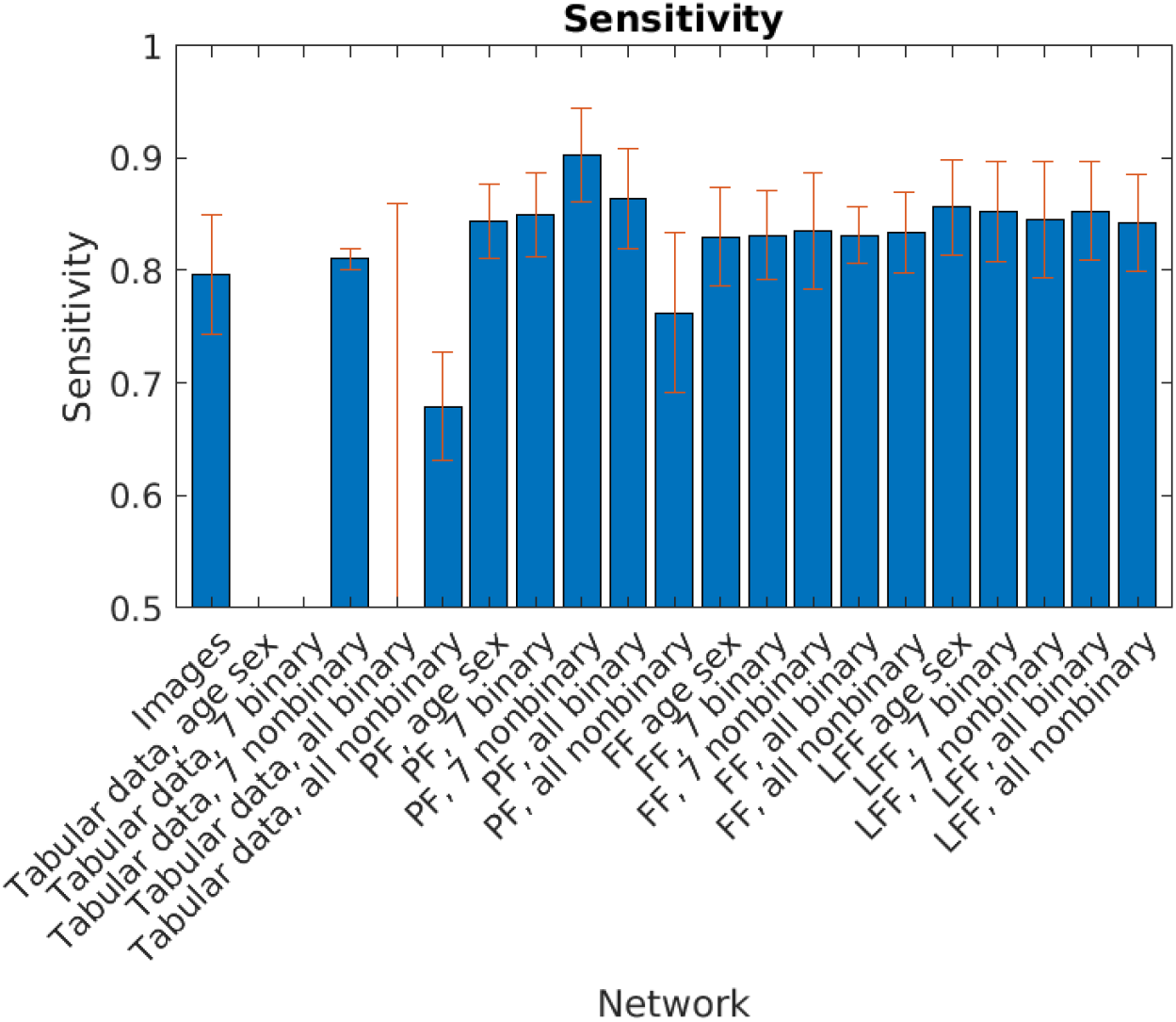
Sensitivity levels assessed for the different networks when using all the available radiographic images per patient. PF, Probability fusion; FF, feature fusion; LFF, learned feature fusion. The error bars represent standard deviations.

**Figure 9.**
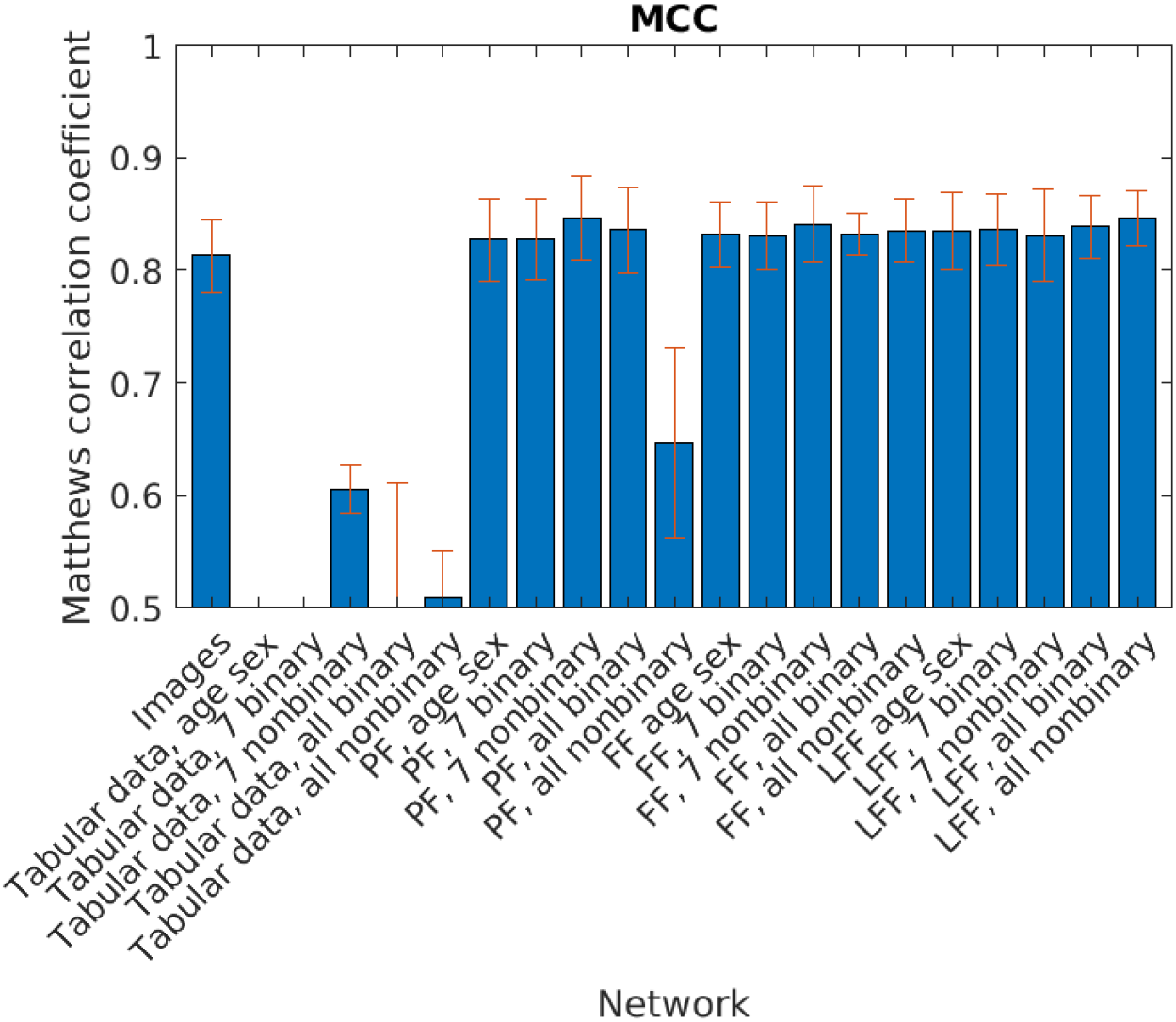
MCC values for the different networks when using all the available radiographic images per patient. PF, Probability fusion; FF, feature fusion; LFF, learned feature fusion. The error bars represent standard deviations.

Our results show that the fusion networks in general improve the AUC, sensitivity and MCC (Table 1). The AUC increased from 0.966 when using only images to 0.987 when using PF and to 0.982 when using LFF (both with seven non-binary variables). Sensitivity increased from 0.796 when using only images, to 0.902 when using PF with seven non-binary variables. MCC increased from 0.813 for image-only classification, to 0.847 when using PF with seven non-binary variables. The average AUC improvements (over all settings and trainings) were small, i.e., 0.5%–2.0%, mainly due to the already high AUC for images only. The average improvements in sensitivity were 6.1% for PF, 4.5% for FF, and 6.7% for LFF. Again, the largest improvement in sensitivity was seen for the seven non-binary variables (8.1%). The average MCC improvement was about 3% for FF and LFF, and the best tabular data were the seven non-binary variables (3.2% improvement). The level of specificity was very similar when using only images or data fusion (0.96–0.98).

### 4.2 Fusion using one or two randomly selected images

The results obtained using a maximum of two randomly selected images per patient are shown in Figures 10–12, and the results for using one randomly selected image per patient are shown in Figures 13–15.

**Figure 10.**
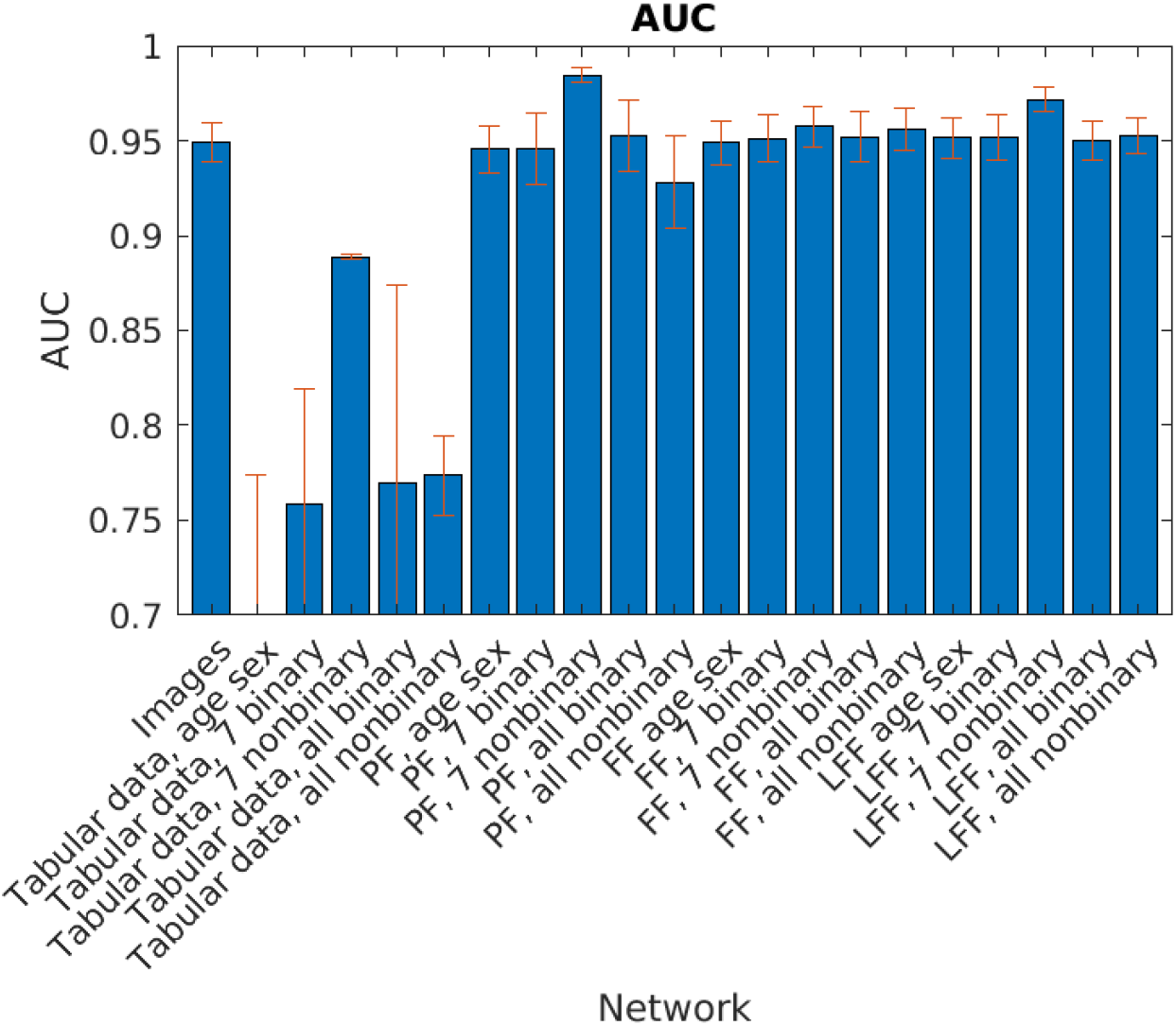
AUC values for the different networks when using a maximum of **two randomly selected images** per patient. PF, Probability fusion; FF, feature fusion; LFF, learned feature fusion. The error bars represent standard deviations.

**Figure 11.**
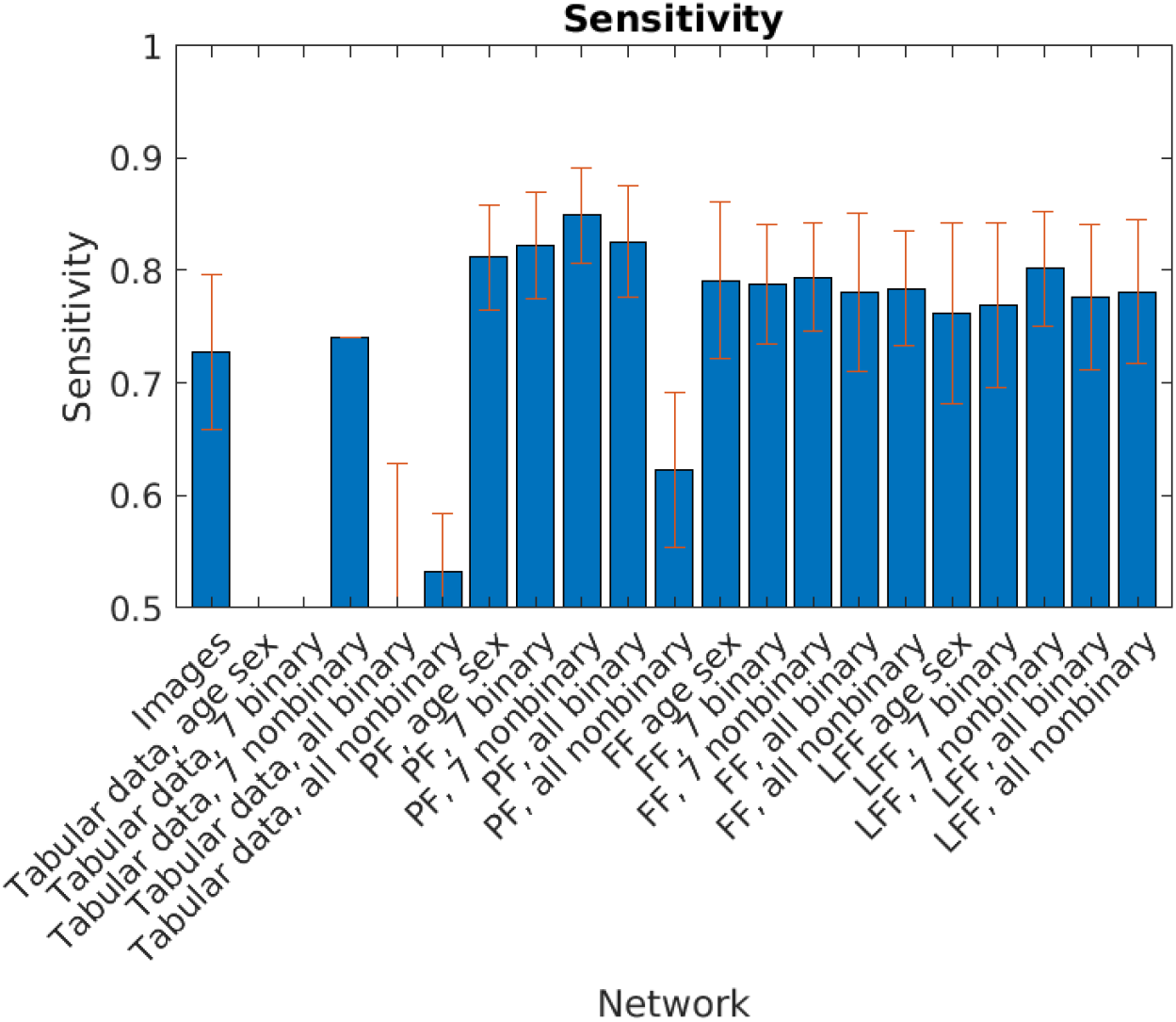
Sensitivity levels for the different networks when using a maximum **of two randomly selected images** per patient. PF, Probability fusion; FF, feature fusion; LFF, learned feature fusion. The error bars represent standard deviations.

**Figure 12.**
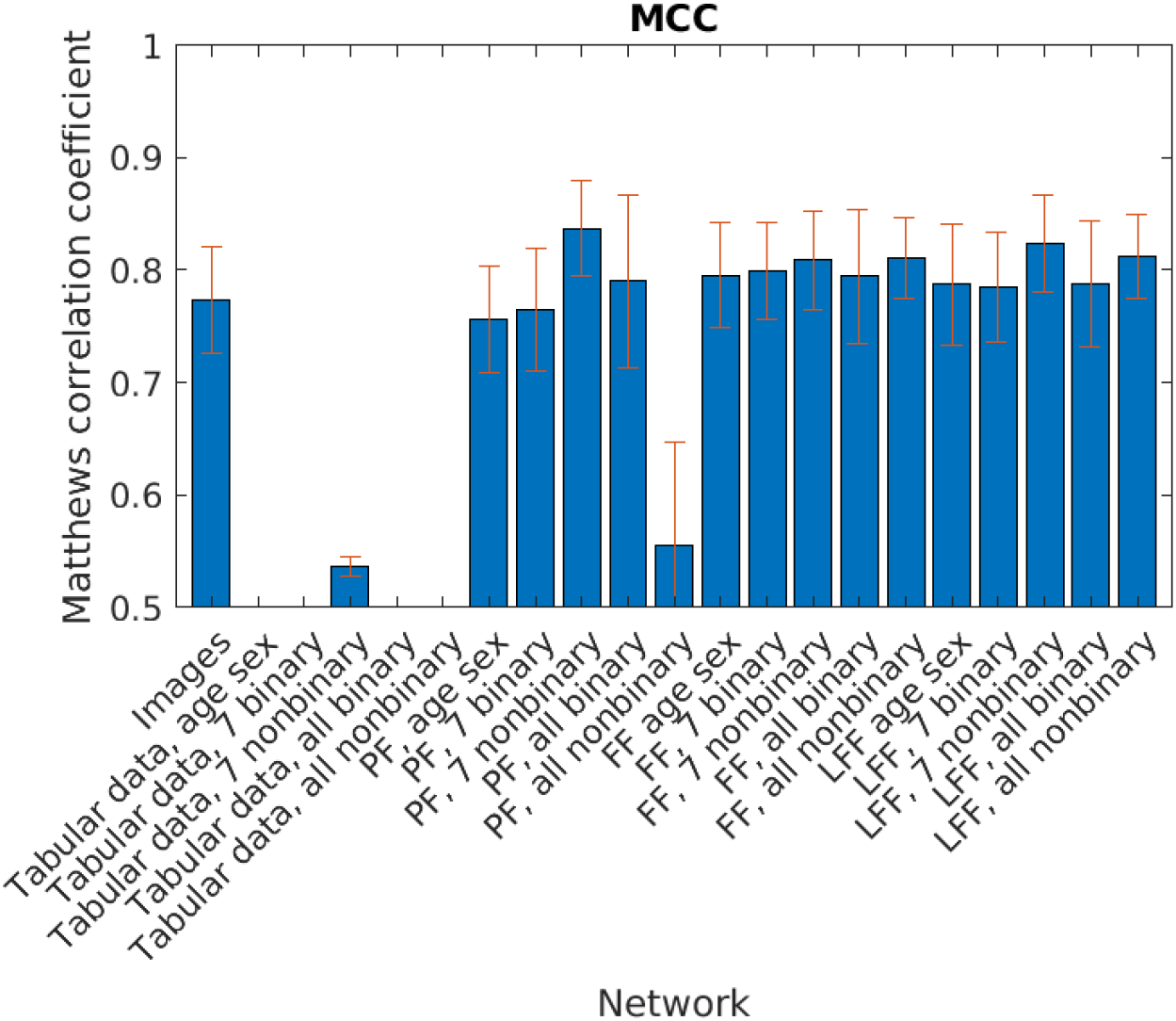
MCC values for the different networks when using a maximum of **two randomly selected images** per patient. PF, Probability fusion; FF, feature fusion; LFF, learned feature fusion. The error bars represent standard deviations.

Using a maximum of two images per patient, the AUC values for the images decreased from 0.966 to 0.949, while PF resulted in an AUC of 0.984 and LFF resulted in an AUC of 0.972 (both with seven non-binary variables), which was close to the results obtained when using all the images. The sensitivity when using only images decreased from 0.796 to 0.724, while the sensitivity for PF with seven non-binary variables decreased from 0.902 to 0.849. The average sensitivity improvements were 8.1% for PF, 8.3% for FF, and 6.9% for LFF. For seven non-binary variables, the improvement in sensitivity was 12.0%.

**Figure 13.**
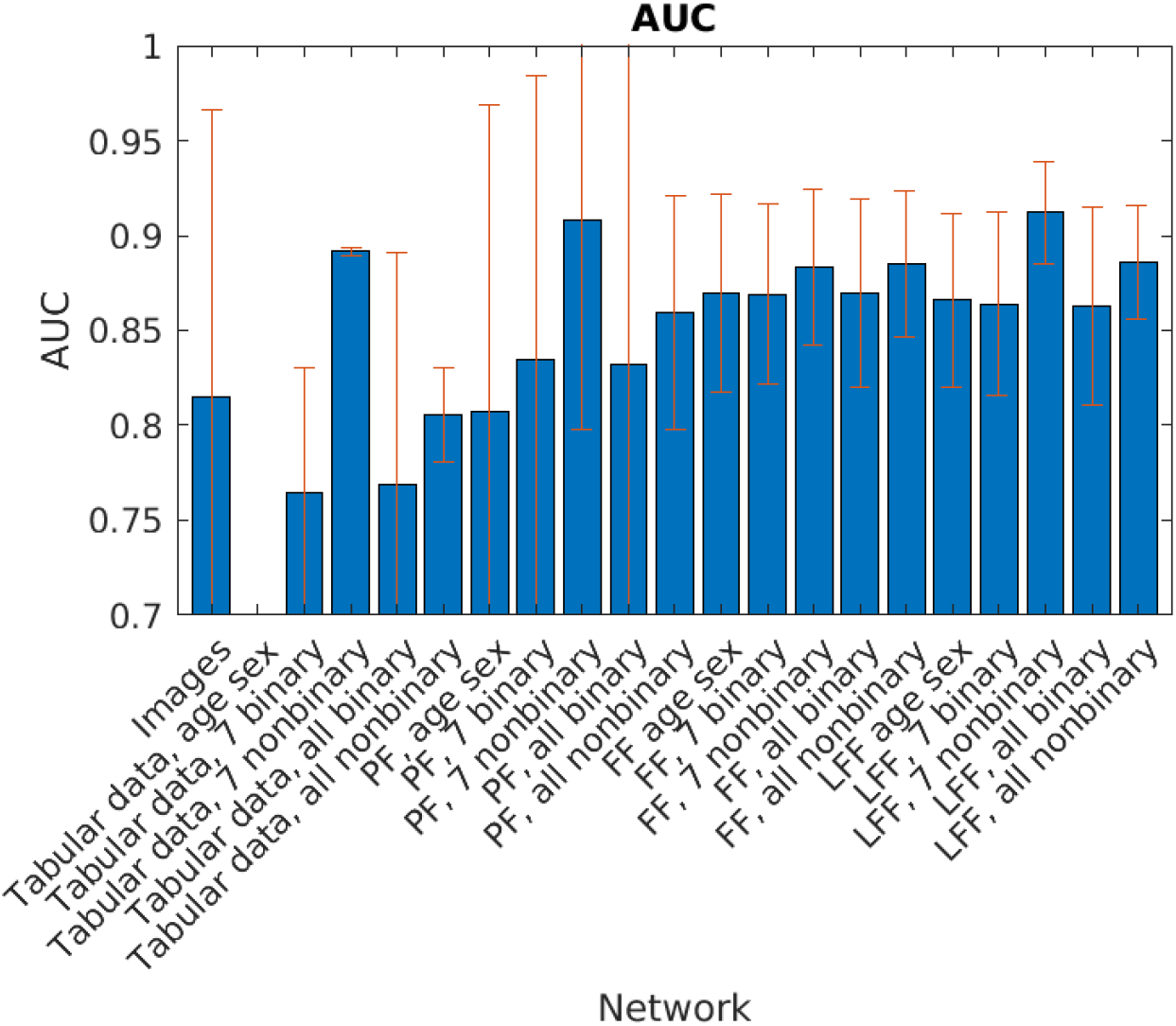
AUC values for the different networks when using **one randomly selected image** per patient. PF, Probability fusion; FF, feature fusion; LFF, learned feature fusion. The error bars represent standard deviations.

**Figure 14.**
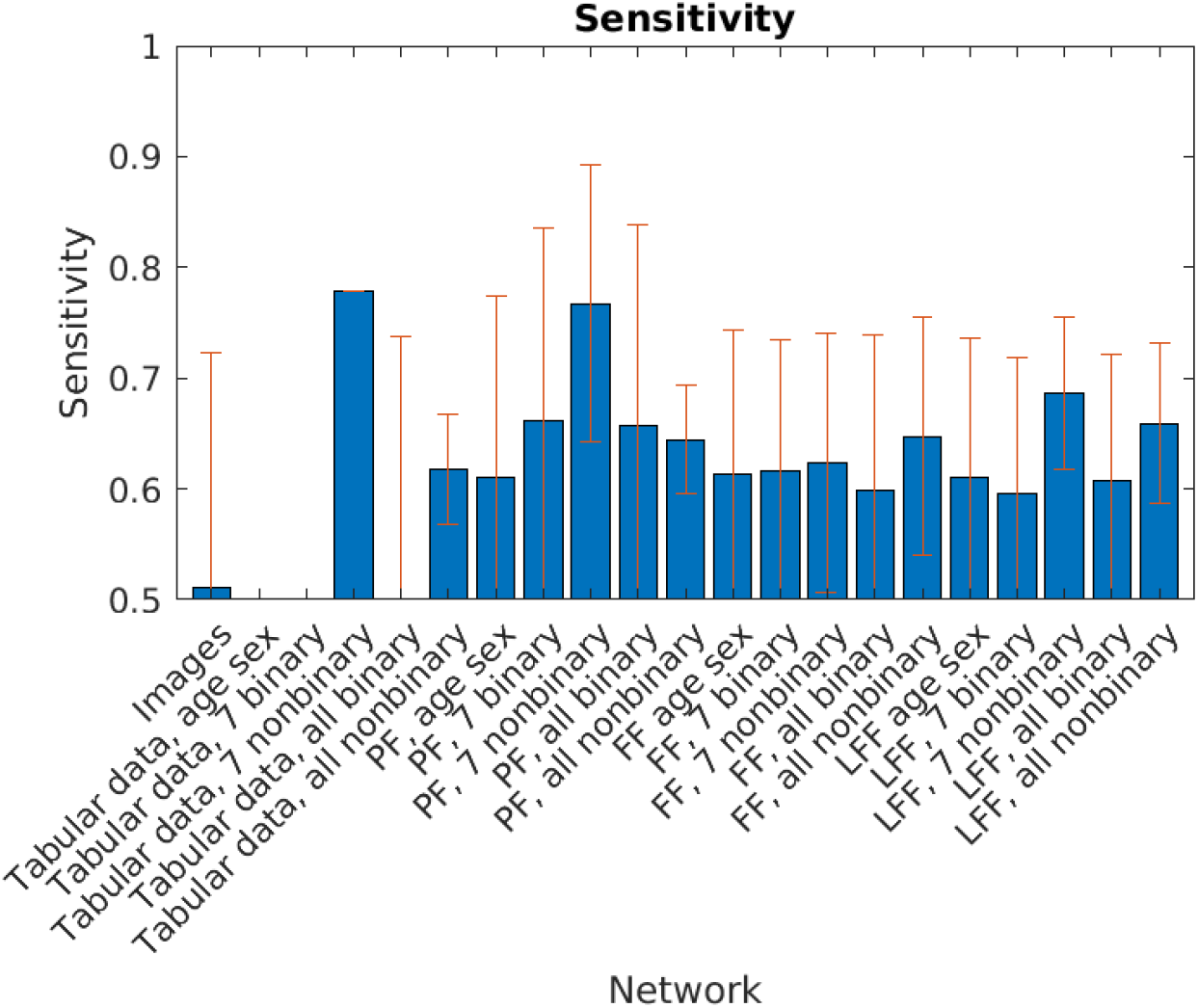
Sensitivity levels for the different networks when using **one randomly selected image** per patient. PF, Probability fusion; FF, feature fusion; LFF, learned feature fusion. The error bars represent standard deviations.

**Figure 15.**
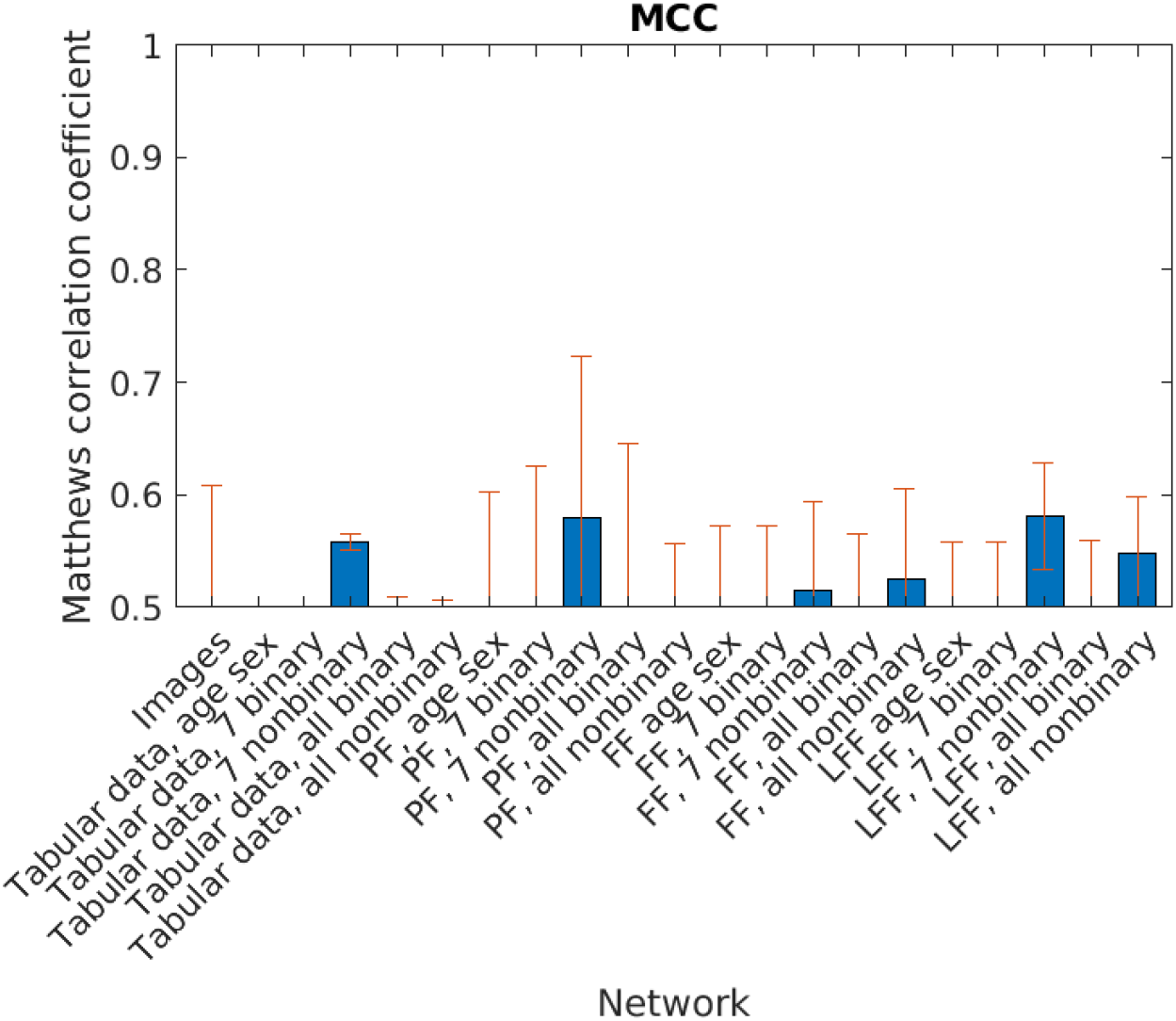
MCC values for the different networks when using **one randomly selected image** per patient. PF, Probability fusion; FF, feature fusion; LFF, learned feature fusion. The error bars represent standard deviations.

Training using one image per patient led to a decrease in the image AUC to 0.815, while PF resulted in an AUC of 0.908 and LFF gave an AUC of 0.912 (both with seven non-binary variables). However, the standard deviation over 25 trainings was much larger for all the networks. When only using images, the sensitivity went down to 0.511, while the sensitivity for PF with seven non-binary variables was 0.767. The average sensitivity improvements were 30.8% for PF, 21.3% for FF, and 23.6% for LFF. However, the sensitivity for fusion was not higher than that seen when only using tabular data.

Taken together, the results show that fusion achieves larger improvements when the different modalities are more balanced, although the estimates were less-precise between the different trainings.

## 5 Discussion

We show that fusing the data from radiographs and electronic health records improves the performances of deep neural networks in discriminating a very rare type of stress fracture (AFF) from normal fractures of the femur (NFF). Although the AUC improvements are not large, going from 0.966 to 0.986, they are clinically relevant considering the importance of avoiding diagnostic errors in medicine. More importantly, the sensitivity increased substantially, from 0.796 to 0.903. These improvements in sensitivity are highly relevant for the clinical situation, where an automated alerting system can attract the attention of the clinician to the very rare event of an AFF. We show that data fusion approaches have greater impacts when the radiographic data and health record data are more balanced. It seems likely that these improvements will be even more pronounced in a more-difficult classification task, such as classifying fractures into several types of fractures.

Fracture classification in orthopedics is popular because it can improve the understanding, diagnosis, and managing of fractures. However, fracture classification systems can be complex, and reproducibility is much poorer compared to the binary classification used in this study. One historical example is the classifying of proximal humerus fractures into fracture types depending on anatomic location and fracture fragment displacement, as described by Neer (Neer, 1970). The classification has been shown to have low inter-observer reliability (50%) and low intra-observer reliability (60%) (Sidor et al., 1993), although it remains as one of the most widely used classification systems. When CNN were applied to the same classification problem, the level of accuracy was very similar to that achieved by human experts, at 65%–86% (Chung et al., 2018), while automated distinction of fractured from non-fractured cases was achieved with almost perfect accuracy (96%). This poor agreement between experts in the assessment of medical conditions is not new but has gained attention recently based on the widely diverging risk assessments made in conjunction with the COVID-19 pandemic and its risks for Society (Ibrahim et al., 2022). Using an arbitrary classification system based on expert opinion, not only limits the accuracy of other human assessments but also that of a mathematical algorithm. In our study, only two types of fractures needed to be discriminated, which might explain the much higher classification accuracy observed compared to other studies.

### 5.1 Comparisons with other studies

Comparisons of our results with the results of previous studies on multimodal fusion in biomedicine are difficult because each multimodal dataset has unique features that can be fused using many different approaches. Furthermore, the results might differ substantially depending on whether or not N-fold CV is used (and M-times repeated N-fold CV is even more uncommon). In a recent review of multimodal fusion (Kline et al., 2022), the mean improvement in predictive accuracy (AUC) was reported to be 6.4% when comparing multimodal and unimodal predictions, which is similar to the improvements seen in our study. In the same review, only 55 out of the 128 reviewed papers were found to have performed N-fold CV. Other studies reported improvements in AUC in the range of 6%–33% for a high number of different multimodal models, when combining up to four different modalities (text, time series, tabular data, and images) (Soenksen et al., 2022) and AUC improvements in the range of 0.02–0.16, irrespective of the approach to fusion that was used (Huang et al., 2020). However, none of these studies have investigated the effect of data fusion on predictive accuracy for fracture classification.

Our study was inspired by a comparison of different fusion approaches to detect breast cancer in MR imaging (Holste et al., 2021). In that study, improvements in AUC from 0.849 to 0.898 were reported when only imaging data were compared with image and non-image features, such as demographics and clinical data. In that study, cross-validation was not performed, which limits the external validity. Few studies have reported on the value of data fusion in musculoskeletal imaging. In the prediction of osteoarthritis progression, an improvement in AUC from 0.76 to 0.80 has been reported, when using radiographs and tabular data such as sex, age, body mass index and radiologic assessment instead of radiologic assessment alone (Tiulpin et al., 2019). In addition, the prediction of mortality for patients with hip fractures was improved from an AUC of 0.717 to 0.786 when combining hip x-ray images, chest x-ray images, and 99 tabular variables (Yenidogan et al., 2021), as compared to a clinical scoring system (Nijmeijer et al., 2016). Similar improvements were obtained (6%) when a unified Holistic AI in a Medicine framework utilized multimodal data from a healthcare system to detect thorax-level spinal fractures (Soenksen et al., 2022). Our results are aligned with these previously reported improvements of multimodal fusion, the main difference being that our performance level is higher when using only imaging data.

### 5.2 Technical limitations

In this work, the high-resolution images were simply zero-padded and down-sampled to 224 × 224 pixels, as the CNN pre-trained on ImageNet expect images of this size. This process removes a lot of information, since the median original size is 2,125 × 2,761 pixels. A better approach could be to train another CNN to first predict the center of each fracture, followed by an automated selection of 224 × 224 or 512 × 512 pixels around the center of the fracture, so as to perform the classification using these patches. It has, for example, been demonstrated that providing attention information for a CNN can improve fracture classification accuracy by some 5%–11% (Liao et al., 2022). However, training a fracture localization CNN on the original high-resolution images is challenging due to the large variations in image size, as well as the substantial computational resources required to perform such a task.

Based on the clinical routine, fracture diagnostics are performed using several radiographic, often orthogonal, projections. This clinical routine provides the clinician with the requisite views, which when analyzed together improve the diagnostic accuracy (Brandser et al., 2000). Compared to this human-based assessment in the clinical routine, our networks only use one image at a time to make a prediction, and then combine the different predictions into a patient-specific prediction. Combining these individual predictions into a single prediction could have a strong impact on network performance. Since we used only one way to combine these predictions, other ways of doing this might yield different results. To assess the effects of different strategies to combine individual predictions, it might be beneficial to use all the images simultaneously. This was not possible in our setting because of the wide variation in the number of images available per patient (varying from 1 to 9). Using Monte Carlo dropout (Gal and Ghahramani, 2016), it would be possible to acquire a measure of the uncertainty for the prediction of each image, or each pairing of imaging and tabular data, potentially utilizing weighted averages of the predictions from each patient, thus limiting the impacts of predictions with high uncertainty.

We used radiographic images obtained from 72 radiology departments in 72 different hospitals throughout Sweden. Consequently, there was substantial variability in terms of the radiology equipment used, patient positioning, education level of the staff, image labeling, and several other factors that might have confounded our results. However, using such a high number of images (4,014) with wide variations in resolution and image quality is beneficial in terms of the generalizability of our results. Initial harmonization of the images could yield even higher accuracy levels, but even without such harmonization the image network was able to reach an AUC of 0.96 when using all the images from each patient.

Since the dataset is unbalanced (15% AFF), it is possible to force the networks to pay more attention to the AFF class in different ways. In this work, we have used only proportional weighting, whereas manually setting the weights could result in a higher level of sensitivity. A simple dense network was used for the tabular data, while other methods, such as random forest and XGBoost, have been shown to outperform deep learning for tabular data (Grinsztajn et al., 2022). In future work, we will test a combination of CNN (or vision transformers) and random forest or XGBoost, to obtain even higher-quality metrics. This could lead to better performance, for example when using all 45 tabular variables, as dense networks have a limited capability to handle uninformative features.

### 5.3 Study limitations

Our results are based on a highly selected sample, comprising almost entirely individuals of Caucasian ethnicity. AFF risk is associated with ethnicity and Asians have a higher risk for AFF, which is linked to increased femoral bow, short stature, and smaller diameter of the femur (Dhanekula et al., 2022). Therefore, the validity of our results for ethnics groups other than Caucasian needs to be evaluated.

Although our dataset contains data from 1,073 patients, a test set of 15% corresponds to 160 patients (about 25 AFF), which may be too small to detect significant differences between the baseline models and fusion models, as well as between different fusion models.

A smaller dataset containing radiographs of NFF and AFF from 373 patients has previously been shared through the AIDA datahub (Hedlund et al., 2020; Zdolsek et al., 2021). The sharing of a combination of radiographs and healthcare data is currently not feasible in European countries due to privacy concerns and regulations.

## Conclusion

In fracture diagnostics, fusion of radiographic and tabular data improves prediction accuracy. The greatest improvement in accuracy achieved in our study related to sensitivity. One of the great advantages associated with automated diagnostic tools is that they can be used in the background to screen large datasets and to alert the clinician to cases requiring specific attention. Specifically, in rare disease patterns such as AFF, deep learning has strong potential to decrease the number of missed diagnoses.

## Data Availability

We have shared our code on Github: https://github.com/wanderine/AFF_fusion . To
share multimodal data is difficult for anonymization purposes.

## Appendix A

The 45 available tabular data variables for each patient are listed in Appendix Table A. The majority of these variables are available as binary or non-binary (n=36). The non-binary version represents the number of days with a certain diagnosis or treatment. The seven pre-selected variables are: age, sex, osteoporosis diagnosis, rheumatoid disease, cortisone treatment, treatment with proton pump inhibitors, and bisphosphonate treatment.

**Table A.**
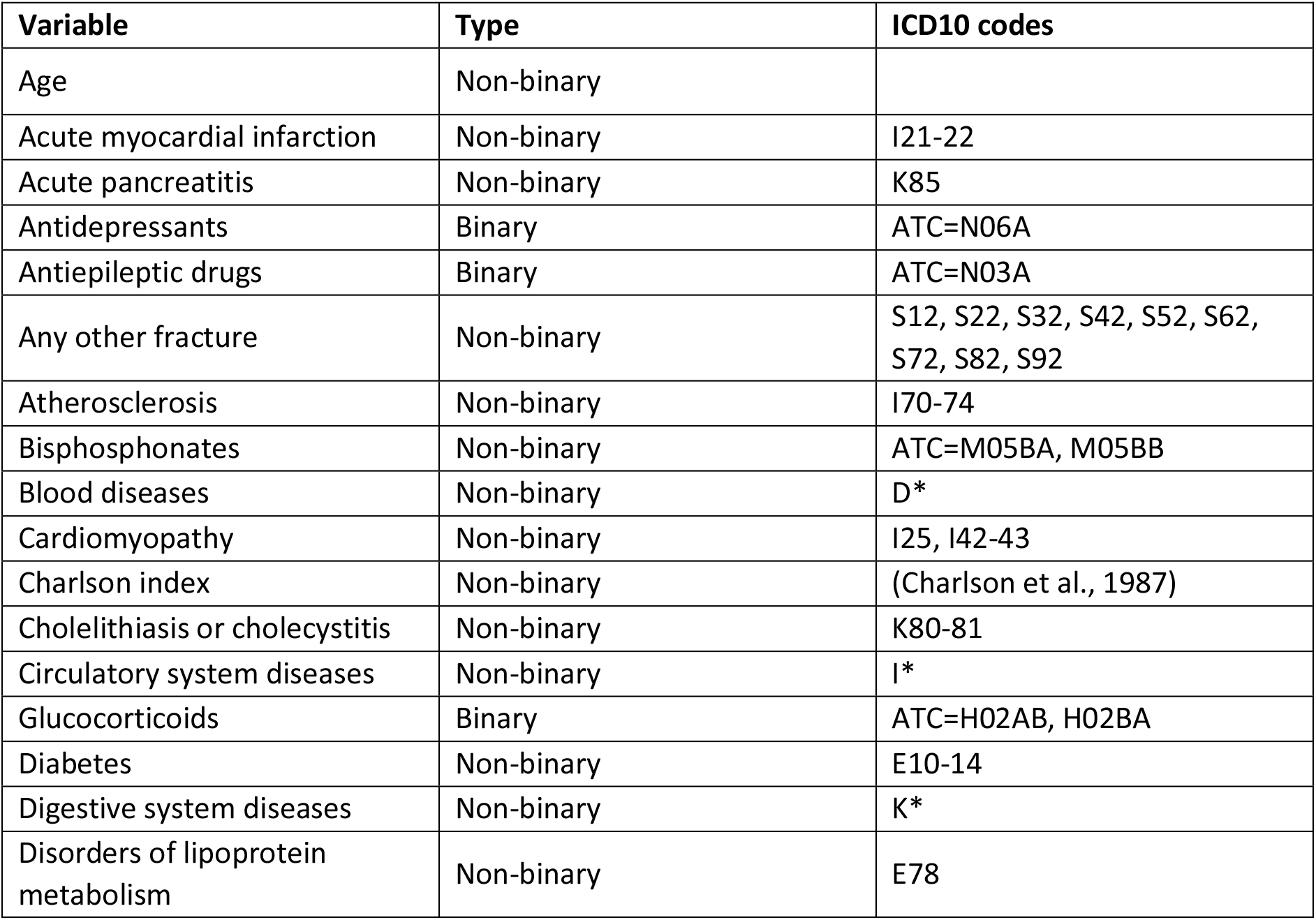
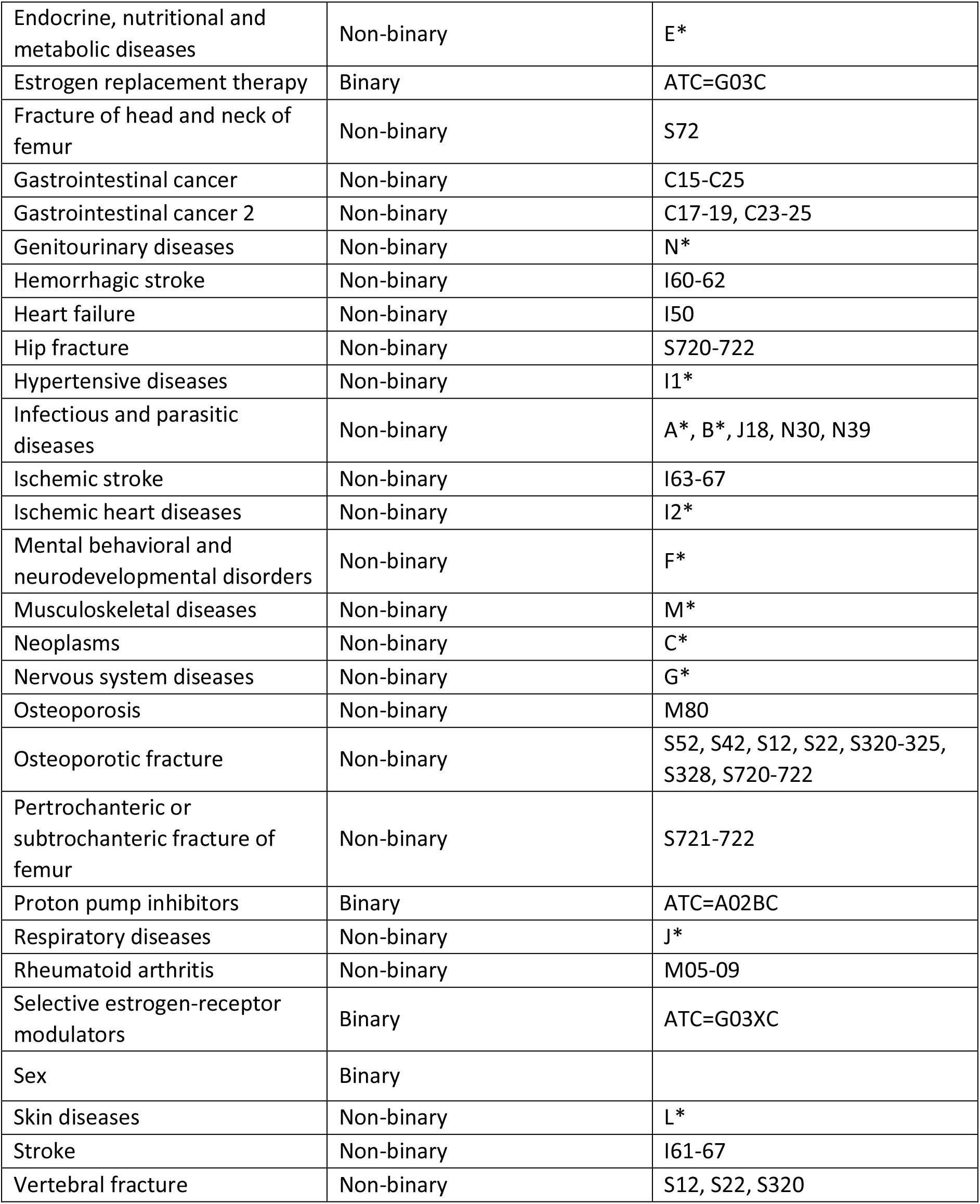
The 45 tabular variables available for each patient, along with their ICD10 codes.

## Ethics

This research study was approved by the Swedish ethical review authority (Dnr. 2021-06482-02).

## Acknowledgments

Anders Eklund was supported by the ITEA/VINNOVA funded project Automation, Surgery Support and Intuitive 3D visualization to optimize the workflow in image-guided therapy (IGT) SysTems (ASSIST) (grant no. 2021-01954). Jörg Schilcher has received ALF funding from Region Östergötland and generous support from the Knut and Alice Wallenberg Foundation. Anders Eklund has previously received graphics hardware from Nvidia.

Alva Nilsson and Oliver Andlid followed the MSc program in biomedical engineering at Linköping University during the period 2018–2022, and parts of this work contributed to their Master’s degree theses.

*Anders Eklund*

Is an Associate Professor at Linköping University. He obtained an MSc in applied physics and electrical engineering in 2007 and a PhD in medical image processing in 2012, both from Linköping University. His research interests include medical image processing, machine learning, statistics, and high-performance computing using graphics cards.

*Jörg Schilcher*

Is an Orthopedic Surgeon at the Department of Orthopedic Surgery, leader of the Lower Extremity Reconstruction Unit, and Professors of orthopedic surgery at Linköping University. Prof. Schilcher’s research focuses on bone healing in the context of fractures and implant integration, specifically primary and revision joint replacement of the hip. He has studied the pathophysiologic, radiologic, and epidemiologic aspects of AFF since 2007. Prof. Schilcher works to improve recognition of atypical femur fractures using machine learning and to identify ways to improve treatment, using specific orthopedic implants and precision medicine to improve the diagnosis and treatment of atypical femur fractures.

## Footnotes

^1^https://www.socialstyrelsen.se/en/statistics-and-data/registers/national-patient-register/

## References

Acosta, J.N., Falcone, G.J., Rajpurkar, P., Topol, E.J., 2022. Multimodal biomedical AI. Nat Med 28, 1773–1784.

Black, D.M., Geiger, E.J., Eastell, R., Vittinghoff, E., Li, B.H., Ryan, D.S., Dell, R.M., Adams, A.L., 2020. Atypical Femur Fracture Risk versus Fragility Fracture Prevention with Bisphosphonates. N Engl J Med 383, 743–753.

Bogl, H.P., Zdolsek, G., Barnisin, L., Moller, M., Schilcher, J., 2022. Surveillance of atypical femoral fractures in a nationwide fracture register. Acta Orthop 93, 229–233.

Bogl, H.P., Zdolsek, G., Michaelsson, K., Hoijer, J., Schilcher, J., 2020. Reduced Risk of Reoperation Using Intramedullary Nailing with Femoral Neck Protection in Low-Energy Femoral Shaft Fractures. The Journal of bone and joint surgery. American volume 102, 1486–1494.

Bradski, G., 2000. The OpenCV library. Dr Dobbs Journal 25, 120–+.

Brandser, E.A., Berbaum, K.S., Dorfman, D.D., Braksiek, R.J., El-Khoury, G.Y., Saltzman, C.L., Marsh, J.L., Clark, W.A., 2000. Contribution of individual projections alone and in combination for radiographic detection of ankle fractures. AJR Am J Roentgenol 174, 1691–1697.

Bruno, M.A., Walker, E.A., Abujudeh, H.H., 2015. Understanding and Confronting Our Mistakes: The Epidemiology of Error in Radiology and Strategies for Error Reduction. Radiographics 35, 1668–1676.

Charlson, M.E., Pompei, P., Ales, K.L., MacKenzie, C.R., 1987. A new method of classifying prognostic comorbidity in longitudinal studies: development and validation. J Chronic Dis 40, 373–383.

Chicco, D., Jurman, G., 2020. The advantages of the Matthews correlation coefficient (MCC) over F1 score and accuracy in binary classification evaluation. BMC Genomics 21, 6.

Chung, S.W., Han, S.S., Lee, J.W., Oh, K.S., Kim, N.R., Yoon, J.P., Kim, J.Y., Moon, S.H., Kwon, J., Lee, H.J., Noh, Y.M., Kim, Y., 2018. Automated detection and classification of the proximal humerus fracture by using deep learning algorithm. Acta Orthop 89, 468–473.

Collaborators, G.B.D.F., 2021. Global, regional, and national burden of bone fractures in 204 countries and territories, 1990-2019: a systematic analysis from the Global Burden of Disease Study 2019. Lancet Healthy Longev 2, e580–e592.

Dell, R.M., Adams, A.L., Greene, D.F., Funahashi, T.T., Silverman, S.L., Eisemon, E.O., Zhou, H., Burchette, R.J., Ott, S.M., 2012. Incidence of atypical nontraumatic diaphyseal fractures of the femur. J Bone Miner Res 27, 2544–2550.

Dhanekula, N.D., Crouch, G., Byth, K., Lau, S.L., Kim, A., Graham, E., Ellis, A., Clifton-Bligh, R.J., Girgis, C.M., 2022. Asian Ethnicity and Femoral Geometry in Atypical Femur Fractures: Independent or Interdependent Risk Factors? JBMR Plus 6, e10607.

Gal, Y., Ghahramani, Z., 2016. Dropout as a bayesian approximation: Representing model uncertainty in deep learning, international conference on machine learning. PMLR, pp. 1050–1059.

Gleadhill, D.N., Thomson, J.Y., Simms, P., 1987. Can more efficient use be made of x ray examinations in the accident and emergency department? Br Med J (Clin Res Ed) 294, 943–947.

Grinsztajn, L., Oyallon, E., Varoquaux, G., 2022. Why do tree-based models still outperform deep learning on tabular data? arXiv preprint arXiv:2207.08815.

Guo, A., Foraker, R.E., MacGregor, R.M., Masood, F.M., Cupps, B.P., Pasque, M.K., 2020. The Use of Synthetic Electronic Health Record Data and Deep Learning to Improve Timing of High-Risk Heart Failure Surgical Intervention by Predicting Proximity to Catastrophic Decompensation. Front Digit Health 2, 576945.

Harborne, K., Hazlehurst, J.M., Shanmugaratnam, H., Pearson, S., Doyle, A., Gittoes, N.J., Choudhary, S., Crowley, R.K., 2016. Compliance with established guidelines for the radiological reporting of atypical femoral fractures. Br J Radiol 89, 20150443.

He, K., Zhang, X., Ren, S., Sun, J., 2016. Deep residual learning for image recognition, Proceedings of the IEEE conference on computer vision and pattern recognition, pp. 770–778.

Hedlund, J., Eklund, A., Lundstrom, C., 2020. Key insights in the AIDA community policy on sharing of clinical imaging data for research in Sweden. Sci Data 7, 331.

Holste, G., Partridge, S.C., Rahbar, H., Biswas, D., Lee, C.I., Alessio, A.M., 2021. End-to-end learning of fused image and non-image features for improved breast cancer classification from MRI, Proceedings of the IEEE/CVF International Conference on Computer Vision, pp. 3294–3303.

Huang, S.C., Pareek, A., Seyyedi, S., Banerjee, I., Lungren, M.P., 2020. Fusion of medical imaging and electronic health records using deep learning: a systematic review and implementation guidelines. NPJ Digit Med 3, 136.

Ibrahim, M.S., Naing, N.N., Abd Aziz, A., Makhtar, M., Mohamed Yusoff, H., Esa, N.K., NI, A.R., Thwe Aung, M.M., Oo, S.S., Ismail, S., Ramli, R.A., 2022. Medical Experts’ Agreement on Risk Assessment Based on All Possible Combinations of the COVID-19 Predictors-A Novel Approach for Public Health Screening and Surveillance. Int J Environ Res Public Health 19.

Kline, A., Wang, H., Li, Y., Dennis, S., Hutch, M., Xu, Z., Wang, F., Cheng, F., Luo, Y., 2022. Multimodal machine learning in precision health: A scoping review. NPJ Digit Med 5, 171.

Kraaijvanger, N., Rijpsma, D., van Leeuwen, H., Edwards, M., 2015. Self-referrals in the emergency department: reasons why patients attend the emergency department without consulting a general practitioner first-a questionnaire study. Int J Emerg Med 8, 46.

Liao, Z., Liao, K., Shen, H., van Boxel, M.F., Prijs, J., Jaarsma, R.L., Doornberg, J.N., Hengel, A.V.D., Verjans, J.W., 2022. CNN Attention Guidance for Improved Orthopedics Radiographic Fracture Classification. IEEE J Biomed Health Inform 26, 3139–3150.

Liu, Y.M., O’Hagan, S., Holdt, F.C., Lahri, S., Pitcher, R.D., 2022. After-hour trauma-radiograph interpretation in the emergency centre of a District Hospital. Afr J Emerg Med 12, 199–207.

Ludvigsson, J.F., Andersson, E., Ekbom, A., Feychting, M., Kim, J.L., Reuterwall, C., Heurgren, M., Olausson, P.O., 2011. External review and validation of the Swedish national inpatient register. BMC Public Health 11, 450.

Matthews, B.W., 1975. Comparison of the predicted and observed secondary structure of T4 phage lysozyme. Biochim Biophys Acta 405, 442–451.

Meier, R.P., Perneger, T.V., Stern, R., Rizzoli, R., Peter, R.E., 2012. Increasing occurrence of atypical femoral fractures associated with bisphosphonate use. Arch Intern Med 172, 930–936.

Neer, C.S., 2nd, 1970. Displaced proximal humeral fractures. I. Classification and evaluation. The Journal of bone and joint surgery. American volume 52, 1077–1089.

Nijmeijer, W.S., Folbert, E.C., Vermeer, M., Slaets, J.P., Hegeman, J.H., 2016. Prediction of early mortality following hip fracture surgery in frail elderly: The Almelo Hip Fracture Score (AHFS). Injury 47, 2138–2143.

Pedregosa, F., Varoquaux, G., Gramfort, A., Michel, V., Thirion, B., Grisel, O., Blondel, M., Prettenhofer, P., Weiss, R., Dubourg, V., Vanderplas, J., Passos, A., Cournapeau, D., Brucher, M., Perrot, M., Duchesnay, E., 2011. Scikit-learn: Machine Learning in Python. Journal of Machine Learning Research 12, 2825–2830.

Pinto, A., Reginelli, A., Pinto, F., Lo Re, G., Midiri, F., Muzj, C., Romano, L., Brunese, L., 2016. Errors in imaging patients in the emergency setting. Br J Radiol 89, 20150914.

Schilcher, J., Koeppen, V., Aspenberg, P., Michaelsson, K., 2014. Risk of atypical femoral fracture during and after bisphosphonate use. N Engl J Med 371, 974–976.

Schilcher, J., Koeppen, V., Aspenberg, P., Michaelsson, K., 2015. Risk of atypical femoral fracture during and after bisphosphonate use. Acta Orthop 86, 100–107.

Schilcher, J., Koeppen, V., Ranstam, J., Skripitz, R., Michaelsson, K., Aspenberg, P., 2013. Atypical femoral fractures are a separate entity, characterized by highly specific radiographic features. A comparison of 59 cases and 218 controls. Bone 52, 389–392.

Schilcher, J., Michaelsson, K., Aspenberg, P., 2011. Bisphosphonate use and atypical fractures of the femoral shaft. N Engl J Med 364, 1728–1737.

Shane, E., Burr, D., Abrahamsen, B., Adler, R.A., Brown, T.D., Cheung, A.M., Cosman, F., Curtis, J.R., Dell, R., Dempster, D.W., Ebeling, P.R., Einhorn, T.A., Genant, H.K., Geusens, P., Klaushofer, K., Lane, J.M., McKiernan, F., McKinney, R., Ng, A., Nieves, J., O’Keefe, R., Papapoulos, S., Howe, T.S., van der Meulen, M.C., Weinstein, R.S., Whyte, M.P., 2014. Atypical subtrochanteric and diaphyseal femoral fractures: second report of a task force of the American Society for Bone and Mineral Research. J Bone Miner Res 29, 1–23.

Shane, E., Burr, D., Ebeling, P.R., Abrahamsen, B., Adler, R.A., Brown, T.D., Cheung, A.M., Cosman, F., Curtis, J.R., Dell, R., Dempster, D., Einhorn, T.A., Genant, H.K., Geusens, P., Klaushofer, K., Koval, K., Lane, J.M., McKiernan, F., McKinney, R., Ng, A., Nieves, J., O’Keefe, R., Papapoulos, S., Sen, H.T., van der Meulen, M.C., Weinstein, R.S., Whyte, M., American Society for, B., Mineral, R., 2010. Atypical subtrochanteric and diaphyseal femoral fractures: report of a task force of the American Society for Bone and Mineral Research. J Bone Miner Res 25, 2267–2294.

Shi, L., Campbell, G., Jones, W.D., Campagne, F., Wen, Z., Walker, S.J., Su, Z., Chu, T.M., Goodsaid, F.M., Pusztai, L., Shaughnessy, J.D., Jr., Oberthuer, A., Thomas, R.S., Paules, R.S., Fielden, M., Barlogie, B., Chen, W., Du, P., Fischer, M., Furlanello, C., Gallas, B.D., Ge, X., Megherbi, D.B., Symmans, W.F., Wang, M.D., Zhang, J., Bitter, H., Brors, B., Bushel, P.R., Bylesjo, M., Chen, M., Cheng, J., Cheng, J., Chou, J., Davison, T.S., Delorenzi, M., Deng, Y., Devanarayan, V., Dix, D.J., Dopazo, J., Dorff, K.C., Elloumi, F., Fan, J., Fan, S., Fan, X., Fang, H., Gonzaludo, N., Hess, K.R., Hong, H., Huan, J., Irizarry, R.A., Judson, R., Juraeva, D., Lababidi, S., Lambert, C.G., Li, L., Li, Y., Li, Z., Lin, S.M., Liu, G., Lobenhofer, E.K., Luo, J., Luo, W., McCall, M.N., Nikolsky, Y., Pennello, G.A., Perkins, R.G., Philip, R., Popovici, V., Price, N.D., Qian, F., Scherer, A., Shi, T., Shi, W., Sung, J., Thierry-Mieg, D., Thierry-Mieg, J., Thodima, V., Trygg, J., Vishnuvajjala, L., Wang, S.J., Wu, J., Wu, Y., Xie, Q., Yousef, W.A., Zhang, L., Zhang, X., Zhong, S., Zhou, Y., Zhu, S., Arasappan, D., Bao, W., Lucas, A.B., Berthold, F., Brennan, R.J., Buness, A., Catalano, J.G., Chang, C., Chen, R., Cheng, Y., Cui, J., Czika, W., Demichelis, F., Deng, X., Dosymbekov, D., Eils, R., Feng, Y., Fostel, J., Fulmer-Smentek, S., Fuscoe, J.C., Gatto, L., Ge, W., Goldstein, D.R., Guo, L., Halbert, D.N., Han, J., Harris, S.C., Hatzis, C., Herman, D., Huang, J., Jensen, R.V., Jiang, R., Johnson, C.D., Jurman, G., Kahlert, Y., Khuder, S.A., Kohl, M., Li, J., Li, L., Li, M., Li, Q.Z., Li, S., Li, Z., Liu, J., Liu, Y., Liu, Z., Meng, L., Madera, M., Martinez-Murillo, F., Medina, I., Meehan, J., Miclaus, K., Moffitt, R.A., Montaner, D., Mukherjee, P., Mulligan, G.J., Neville, P., Nikolskaya, T., Ning, B., Page, G.P., Parker, J., Parry, R.M., Peng, X., Peterson, R.L., Phan, J.H., Quanz, B., Ren, Y., Riccadonna, S., Roter, A.H., Samuelson, F.W., Schumacher, M.M., Shambaugh, J.D., Shi, Q., Shippy, R., Si, S., Smalter, A., Sotiriou, C., Soukup, M., Staedtler, F., Steiner, G., Stokes, T.H., Sun, Q., Tan, P.Y., Tang, R., Tezak, Z., Thorn, B., Tsyganova, M., Turpaz, Y., Vega, S.C., Visintainer, R., von Frese, J., Wang, C., Wang, E., Wang, J., Wang, W., Westermann, F., Willey, J.C., Woods, M., Wu, S., Xiao, N., Xu, J., Xu, L., Yang, L., Zeng, X., Zhang, J., Zhang, L., Zhang, M., Zhao, C., Puri, R.K., Scherf, U., Tong, W., Wolfinger, R.D., Consortium, M., 2010. The MicroArray Quality Control (MAQC)-II study of common practices for the development and validation of microarray-based predictive models. Nat Biotechnol 28, 827–838.

Sidor, M.L., Zuckerman, J.D., Lyon, T., Koval, K., Cuomo, F., Schoenberg, N., 1993. The Neer classification system for proximal humeral fractures. An assessment of interobserver reliability and intraobserver reproducibility. The Journal of bone and joint surgery. American volume 75, 1745–1750.

Soenksen, L.R., Ma, Y., Zeng, C., Boussioux, L., Villalobos Carballo, K., Na, L., Wiberg, H.M., Li, M.L., Fuentes, I., Bertsimas, D., 2022. Integrated multimodal artificial intelligence framework for healthcare applications. NPJ Digit Med 5, 149.

Starr, J., Tay, Y.K.D., Shane, E., 2018. Current Understanding of Epidemiology, Pathophysiology, and Management of Atypical Femur Fractures. Curr Osteoporos Rep 16, 519–529.

Tampu, I.E., Eklund, A., Haj-Hosseini, N., 2022. Inflation of test accuracy due to data leakage in deep learning-based classification of OCT images. Scientific Data 9, 580.

Tiulpin, A., Klein, S., Bierma-Zeinstra, S.M.A., Thevenot, J., Rahtu, E., Meurs, J.V., Oei, E.H.G., Saarakkala, S., 2019. Multimodal Machine Learning-based Knee Osteoarthritis Progression Prediction from Plain Radiographs and Clinical Data. Sci Rep 9, 20038.

Welfare, S.N.B.o.H.a., 2020. Analys av hur patienter besöker somatiska akutmottagningar under och efter första covid-19-vågen.

Whang, J.S., Baker, S.R., Patel, R., Luk, L., Castro, A., 3rd, 2013. The causes of medical malpractice suits against radiologists in the United States. Radiology 266, 548–554.

Willis, B.H., 2012. Empirical evidence that disease prevalence may affect the performance of diagnostic tests with an implicit threshold: a cross-sectional study. BMJ Open 2, e000746.

Yenidogan, B., Pathak, S., Geerdink, J., Hegeman, J.H., Van Keulen, M., 2021. Multimodal machine learning for 30-days post-operative mortality prediction of elderly hip fracture patients., International Conference on Data Mining Workshops.

Zdolsek, G., Chen, Y., Bogl, H.P., Wang, C., Woisetschlager, M., Schilcher, J., 2021. Deep neural networks with promising diagnostic accuracy for the classification of atypical femoral fractures. Acta Orthop 92, 394–400.

